# Vitamin D Status and Supplementation in Patients Undergoing Spine Surgery: A Systematic Review and Meta-analysis

**DOI:** 10.64898/2026.08.02.26359512

**Authors:** Farzan Fahim, Fatemeh Vosoughian, Amirmahdi Mojtahedzadeh, Mahla Rakhshani, Barbod Mahdavi, Kiana Taghipoor, Rauf Rostami, Reihane Qahremani, Fatemeh Gheibi, Fatemeh Deldar, Ramtin Shemshadigolafzani, Saeed-Rezaali, Nahal Badavi, Pardis Fathabadi, Alireza Zali

## Abstract

**Background:** Vitamin D may influence bone healing and recovery after spine surgery, but its clinical effects remain uncertain.

**Objective:** To evaluate clinical, functional, and bone-related outcomes associated with vitamin D status and supplementation in patients undergoing spine surgery.

**Methods:** This systematic review and meta-analysis was registered in PROSPERO (CRD420261321165) and reported according to PRISMA 2020. PubMed/MEDLINE, Scopus, Web of Science, Embase, and the Cochrane Library were searched from inception through 15 January 2026. Random-effects meta-analyses used mean differences (MDs) for continuous outcomes and risk ratios (RRs) for dichotomous outcomes. Direct supplementation analyses were prioritized; analyses combining supplementation, baseline vitamin D status, or co-interventions were exploratory. Risk of bias was assessed using design-specific Joanna Briggs Institute tools.

**Results:** Thirteen studies were included. Four supplementation studies involving 177 participants showed no significant reduction in postoperative pain (MD −0.82, 95% CI −2.50 to 0.86; I² = 84.3%). Three studies involving 277 participants showed lower Oswestry Disability Index scores in an exploratory analysis (MD −6.40, 95% CI −10.41 to −2.39; I² = 96.8%). Three supplementation studies involving 128 participants showed no significant improvement in fusion rate (RR 1.17, 95% CI 0.78–1.74; I² = 31.5%). An exploratory four-study fusion analysis was also nonsignificant (RR 1.24, 95% CI 0.98–1.57; I² = 45.5%).

**Conclusion:** Current evidence does not establish that vitamin D supplementation independently improves postoperative pain or fusion rates after spine surgery. Possible benefits for disability and other bone-related outcomes remain uncertain because of small study numbers, heterogeneity, co-interventions, and residual confounding. Larger randomized trials stratified by baseline vitamin D status are needed.

## Introduction

Osteoporosis is a major global health problem affecting more than 200 million individuals worldwide, with a particularly high prevalence among postmenopausal women and older adults [1,2]. Its burden is expected to increase as populations age, accompanied by a growing incidence of fragility fractures and an increasing number of older patients requiring spinal reconstruction or fusion procedures [3,4]. Vertebral fractures are among the most clinically important manifestations of osteoporosis and may result in persistent pain, deformity, impaired mobility, neurological compromise, and, in selected patients, the need for surgical intervention [5,6]. More broadly, reduced bone quality in patients undergoing spine surgery may increase the risk of implant loosening, cage subsidence, delayed fusion, pseudarthrosis, junctional complications, and revision surgery [7,8].

Successful spinal arthrodesis depends on coordinated osteogenesis, graft incorporation, mineralization, and skeletal remodeling across the fusion bed. These processes may be impaired in patients with reduced bone mineral density, altered bone turnover, or inadequate calcium and vitamin D availability [9,10]. Vitamin D contributes to intestinal calcium absorption, regulation of parathyroid hormone secretion, osteoblast differentiation, osteoclast signaling, and maintenance of skeletal and neuromuscular function [11,12]. Deficiency may therefore create an unfavorable biological environment for bone healing and potentially contribute to delayed arthrodesis or pseudarthrosis [13]. Such complications may prolong recovery, increase postoperative pain and disability, expose instrumentation to sustained mechanical stress, and contribute to fracture or reoperation [14,15].

Perioperative optimization of bone health may include nutritional intervention, physical activity, calcium and vitamin D replacement, and pharmacological treatment of osteoporosis [16,17]. Calcium and vitamin D supplementation have shown variable effects on bone mineral density and fracture prevention, with outcomes influenced by baseline nutritional status, adherence, concurrent treatment, and the population studied [18,19]. Other anti-osteoporosis agents, including bisphosphonates, denosumab, and anabolic therapies, may also affect bone density, implant stability, subsidence, fracture risk, and fusion-related outcomes; however, their effects depend on treatment selection, timing, dose, and the severity of underlying skeletal disease [20].

Despite a strong biological rationale for identifying and correcting vitamin D deficiency, the clinical relevance of vitamin D status and supplementation in patients undergoing spine surgery remains uncertain. Available studies differ substantially in surgical procedure, baseline vitamin D concentration, supplementation formulation and dose, comparator definition, concurrent anti-osteoporosis treatment, follow-up duration, and the methods used to assess pain, disability, fusion, bone quality, and implant-related complications. Some studies have directly evaluated vitamin D-containing supplementation, whereas others have examined outcomes according to baseline vitamin D status, supplementation dose, or broader bone-metabolic characteristics. These related but clinically distinct comparisons have produced inconsistent findings and complicate interpretation of the existing evidence.

Accordingly, this systematic review and meta-analysis aimed to evaluate the association of vitamin D status and vitamin D-containing supplementation with postoperative pain, disability, radiographic fusion, bone quality, implant-related complications, fractures, functional recovery, and safety in patients undergoing spine surgery. Pain measured using the Visual Analog Scale was designated as the primary quantitative outcome, while disability measured using the Oswestry Disability Index and radiographic fusion rate were evaluated as secondary outcomes.

## Methods

### Protocol and reporting

This systematic review and meta-analysis was conducted according to a predefined protocol and reported in accordance with the Preferred Reporting Items for Systematic Reviews and Meta-Analyses 2020 statement. The completed PRISMA 2020 checklist is provided in Supplementary Material 6. The review was registered in the International Prospective Register of Systematic Reviews (PROSPERO; registration number CRD420261321165). The protocol specified the review question, eligibility criteria, information sources, study-selection procedures, data extraction, risk-of-bias assessment, outcomes, quantitative synthesis, subgroup analyses, and sensitivity analyses.

### Search strategy and information sources

A systematic search was conducted in PubMed/MEDLINE, Scopus, Web of Science, Embase, and the Cochrane Library from database inception through 15 January 2026. No restrictions were imposed on publication year or language. Reports published in languages other than English were assessed using machine-assisted translation when required.

The search strategies combined controlled vocabulary and free-text terms related to spine surgery, spinal fusion, vitamin D, calcium-vitamin D supplementation, vitamin D analogues, bone healing, pain, disability, fusion, bone mineral density, implant stability, and postoperative complications. Spine-related terms included “spinal surgery,” “spine surgery,” “spinal fusion,” “spondylodesis,” “spinal fixation,” “lumbar fusion,” “posterior lumbar interbody fusion,” “transforaminal lumbar interbody fusion,” “anterior lumbar interbody fusion,” and “instrumented fusion.” Exposure-related terms included “vitamin D,” “vitamin D3,” “cholecalciferol,” “calciferol,” “calcitriol,” “1,25-dihydroxyvitamin D3,” “alfacalcidol,” “25-hydroxyvitamin D,” “calcium,” “calcium carbonate,” and “calcium citrate.” Outcome-related terms included “fusion rate,” “pseudarthrosis,” “nonunion,” “bone healing,” “bone mineral density,” “implant loosening,” “cage subsidence,” “revision surgery,” “Visual Analog Scale,” and “Oswestry Disability Index.”

Search syntax was adapted to the indexing system and interface of each database. The complete database-specific search strategies are provided in Supplementary Material 1.

### Eligibility criteria

Eligibility criteria were defined according to the population, intervention or exposure, comparator, outcomes, and study-design framework.

### Population

Eligible populations included human participants undergoing cervical, thoracic, lumbar, lumbosacral, or thoracolumbar spine surgery. Surgical procedures included decompression, instrumented stabilization, posterolateral fusion, posterior lumbar interbody fusion, transforaminal lumbar interbody fusion, anterior or lateral lumbar interbody fusion, anterior cervical interbody fusion, deformity correction, and other spinal fusion or fixation procedures.

### Intervention or exposure

Eligible interventions and exposures included vitamin D supplementation, calcium–vitamin D supplementation, cholecalciferol, activated vitamin D analogues such as calcitriol or alfacalcidol, and other perioperative treatment regimens containing vitamin D. Studies evaluating baseline vitamin D status, vitamin D supplementation dose, or correction of vitamin D deficiency in relation to postoperative outcomes were also eligible.

Studies involving concurrent anti-osteoporosis treatment were retained when the vitamin D-related exposure or treatment group could be identified. Comparisons involving additional active treatments, such as denosumab, bisphosphonates, or other bone-modifying agents, were considered clinically distinct and were interpreted cautiously because the independent contribution of vitamin D could not always be isolated.

### Comparator

Eligible comparators included placebo, routine care, no supplementation, calcium alone, an alternative anti-osteoporosis treatment, a different vitamin D dose, untreated vitamin D deficiency, or a comparison group defined according to baseline vitamin D or bone-metabolic status. A formal comparator was not required for narrative inclusion when an eligible study reported clinically relevant postoperative outcomes associated with vitamin D treatment or status.

### Outcomes

The primary quantitative outcome was postoperative pain measured using the Visual Analog Scale. Secondary quantitative outcomes were disability measured using the Oswestry Disability Index and radiographic fusion rate.

Additional eligible outcomes included time to fusion, nonunion or pseudarthrosis, bone mineral density, computed-tomography-derived bone quality, implant loosening, cage migration or subsidence, proximal junctional kyphosis or failure, vertebral or sacral fracture, revision surgery, quality of life, balance, postural stability, inflammatory markers, muscle-related outcomes, and adverse events.

### Study design

Eligible study designs included randomized controlled trials, nonrandomized controlled or interventional studies, prospective and retrospective cohort studies, case-control studies, before– after studies, and case series including at least 10 participants.

Reviews, meta-analyses, editorials, commentaries, case reports, nonhuman studies, in vitro investigations, conference abstracts without sufficient extractable data, and case series including fewer than 10 participants were excluded. Studies were also excluded when they did not involve spine surgery, did not evaluate vitamin D status or a vitamin D-containing intervention, or lacked relevant and extractable clinical outcome data.

Studies evaluating calcium, phosphate, the calcium–phosphate product, or parathyroid hormone without evaluating vitamin D status or a vitamin D-containing intervention were not eligible for inclusion.

### Study selection

All records retrieved from the database searches were imported into EndNote and Rayyan, and duplicate records were removed before screening. Four reviewers (B.M., K.T., F.D., and R.Sh.) participated in title and abstract screening according to the predefined eligibility criteria. Potentially eligible or uncertain records were retained for full-text assessment. Screening conflicts were discussed and resolved through consensus, and reasons for exclusion were documented.

Full-text eligibility assessment was conducted by two reviewers (R.R. and K.T.). Each report was classified as included or excluded, and a predefined reason was recorded for every full-text exclusion. The exclusion and inclusion forms were designed by F.V. For excluded studies, the title, digital object identifier, country of origin, screening decision, and reason for exclusion were recorded. For included studies, the title, digital object identifier, study design, population, vitamin D or bone-status subgroup, intervention, comparator, and reported outcomes were documented.

The finalized full-text screening workbook, including decisions for all 154 assessed reports, the final 13-study evidence set, and the 141 exclusions grouped into the three PRISMA categories, is provided in Supplementary Material 2. Conflicting or incomplete entries in the source screening files were adjudicated against the prespecified PICOS criteria. The overall study-selection process was documented using a PRISMA 2020 flow diagram.

### Data extraction

Data were extracted using a standardized form designed by F.F. and F.V. Two reviewers (R.R. and F.D.) independently extracted the available data. Disagreements or uncertainties were resolved through discussion and consensus.

Extracted study-level variables included first author, publication year, digital object identifier, country, participating center or centers, study design, study period, eligibility criteria, and funding source. Participant-level variables included sample size, age, sex, body mass index, smoking status, comorbidities, baseline serum vitamin D concentration, bone mineral density, T-score, and the proportion of participants with osteopenia or osteoporosis.

Surgical variables included spinal region, surgical indication, surgical procedure, discectomy, instrumentation, cage implantation, number and dimensions of screws, number of rods, bone cement augmentation, expandable screws, and other relevant technical characteristics. Information concerning medications that could affect bone metabolism, including corticosteroids, antiepileptic medications, warfarin, bisphosphonates, denosumab, and other anti-osteoporosis agents, was also extracted.

Vitamin D-related variables included baseline serum 25-hydroxyvitamin D concentration, intervention type, vitamin D formulation, route of administration, dosage, treatment timing, treatment duration, calcium coadministration, concurrent bone-modifying treatment, and follow-up serum vitamin D concentration.

Extracted outcomes included pain, disability, fusion rate, time to fusion, nonunion or pseudarthrosis, bone mineral density, computed-tomography-derived bone quality, implant loosening, cage migration or subsidence, proximal junctional complications, vertebral or sacral fracture, adjacent-segment degeneration, revision surgery, quality of life, balance, postural stability, inflammatory markers, muscle-related biomarkers, and adverse events. The complete extraction dataset is provided in Supplementary Material 3.

### Outcome definitions and analytical hierarchy

Pain intensity measured using the Visual Analog Scale was designated as the primary quantitative outcome. Disability measured using the Oswestry Disability Index and radiographic fusion rate were designated as secondary quantitative outcomes.

For continuous outcomes, group-level means, standard deviations, and sample sizes were extracted at the postoperative time point selected for the corresponding analysis. Lower mean differences favored the vitamin D-related group for the Visual Analog Scale and Oswestry Disability Index outcomes. When multiple eligible follow-up assessments were reported, the selected time point was applied consistently within the relevant analysis, and alternative eligible time points were examined in sensitivity analyses when appropriate.

For fusion rate, the number of participants achieving radiographic fusion and the corresponding group denominator were extracted. Risk ratios greater than 1 favored the vitamin D-related group. Fusion definitions were retained as reported by the original investigators and included assessments based on plain radiography, computed tomography, or combined imaging methods. Studies directly comparing vitamin D supplementation or a vitamin D-containing treatment with a control condition were distinguished from studies comparing baseline vitamin D status, supplementation dose, bone-metabolic status, or treatment regimens involving major co-interventions. Intervention-focused analyses were prioritized. Analyses combining supplementation with baseline-status or co-intervention comparisons were regarded as exploratory because these comparisons did not estimate an identical treatment effect.

The principal Visual Analog Scale analysis was therefore restricted to direct supplementation studies. A broader analysis incorporating a clinically distinct bone-status and co-intervention comparison was conducted as an exploratory analysis. Similarly, the principal fusion-rate analysis was restricted to supplementation or treatment studies, whereas the four-study analysis incorporating a clinically distinct bone-status comparison was considered exploratory. The Oswestry Disability Index analysis was also regarded as exploratory because it combined direct supplementation with vitamin D or bone-status comparisons.

### Risk-of-bias assessment

Two reviewers (M.R. and R.Q.) independently assessed methodological quality using the Joanna Briggs Institute critical appraisal tool appropriate to each study design. Randomized controlled trials were evaluated using the JBI randomized controlled trial checklist, cohort studies using the JBI cohort checklist, case-control studies using the JBI case-control checklist, and nonrandomized interventional studies using the corresponding JBI quasi-experimental checklist. Each item was judged as “Yes,” “No,” “Unclear,” or “Not applicable.” Overall study-level judgments were categorized as low, moderate, or high risk of bias according to the prespecified decision framework used by the review team. Disagreements were resolved through discussion and consensus. Summary study-level findings are reported in the Results, and the complete item-level assessments are provided in Supplementary Material 4.

### Data synthesis and quantitative analysis

Meta-analyses were performed using R version 4.5.1. Continuous outcomes were synthesized as mean differences with 95% confidence intervals, whereas dichotomous outcomes were synthesized as risk ratios with 95% confidence intervals.

Random-effects models were selected a priori because clinical and methodological heterogeneity was anticipated across patient populations, baseline vitamin D status, supplementation regimens, comparator definitions, surgical procedures, concurrent anti-osteoporosis treatments, outcome definitions, imaging methods, follow-up durations, and assessment time points. Between-study variance was estimated using restricted maximum likelihood. Random-effects estimates were considered the principal results, while common-effect estimates were regarded as supplementary.

Statistical heterogeneity was assessed using Cochran’s Q test and quantified using I², τ, and τ². I² was interpreted as the proportion of total variability attributable to between-study heterogeneity rather than sampling error. Prediction intervals were calculated when estimable to describe the range of effects that might be expected in a future comparable study.

For multi-arm studies, only groups relevant to the specified comparison were included. Shared comparator groups were not entered more than once within the same pooled analysis. When a study contained clinically distinct treatment groups or major co-interventions, the comparison was analyzed separately or incorporated only into an exploratory analysis.

### Subgroup, sensitivity, and influence analyses

Leave-one-out sensitivity analyses were performed by repeating each meta-analysis after sequential omission of one study to determine whether the pooled estimate or heterogeneity was disproportionately influenced by an individual study.

For the Visual Analog Scale outcome, the intervention-focused analysis excluded Shi et al. because its groups were defined by baseline bone status and included denosumab as a co-intervention. A broader five-study analysis was retained as exploratory. An additional sensitivity analysis excluded Haddadi et al. because influence diagnostics indicated a disproportionate effect on the pooled estimate. Influence analysis was performed using squared Pearson residuals and standard influence diagnostics.

For fusion rate, subgroup analysis was performed according to comparison type, separating direct supplementation or treatment studies from the clinically distinct bone-status and co-intervention comparison. A supplementation-only analysis was conducted as the principal intervention-focused synthesis, while a broader four-study analysis was retained as exploratory. Leave-one-out analyses were conducted for both the complete fusion dataset and the supplementation-only subgroup. An additional sensitivity analysis used the six-month fusion data reported by Xu et al. to evaluate whether the pooled estimate was dependent on the selected follow-up time.

The exploratory analyses, subgroup analyses, influence diagnostics, leave-one-out analyses, and follow-up-time sensitivity analyses are provided in Supplementary Material 5.

### Small-study effects

Funnel plots were generated to explore potential small-study effects for the Visual Analog Scale, Oswestry Disability Index, exploratory overall fusion-rate, and supplementation-only fusion-rate analyses. Trim-and-fill analysis was also performed for the Visual Analog Scale outcome as an exploratory assessment.

Because fewer than 10 studies contributed to each quantitative outcome, funnel-plot interpretation and trim-and-fill findings were considered unreliable for establishing publication bias. These analyses were therefore treated as exploratory, and no firm conclusion regarding publication bias or small-study effects was based on them. The funnel plots and trim-and-fill analysis are provided in Supplementary Material 5.

## Results

### Study selection

The database searches identified 3,793 records. After removal of 1,484 duplicate records, 2,309 records underwent title and abstract screening. Of these, 2,155 were excluded because they did not satisfy the predefined eligibility criteria, leaving 154 reports for full-text assessment.

Following full-text review, 141 reports were excluded: 16 because of an ineligible study design, 56 because they did not report suitable outcomes, and 69 because they did not evaluate an eligible intervention or exposure. Thirteen studies met the eligibility criteria for the systematic review, and seven provided sufficiently compatible data to contribute to at least one quantitative synthesis. The finalized decisions for all 154 full-text reports are provided in Supplementary Material 2. The study-selection process is presented in the PRISMA 2020 flow diagram (Figure 1).

**Figure 1.**
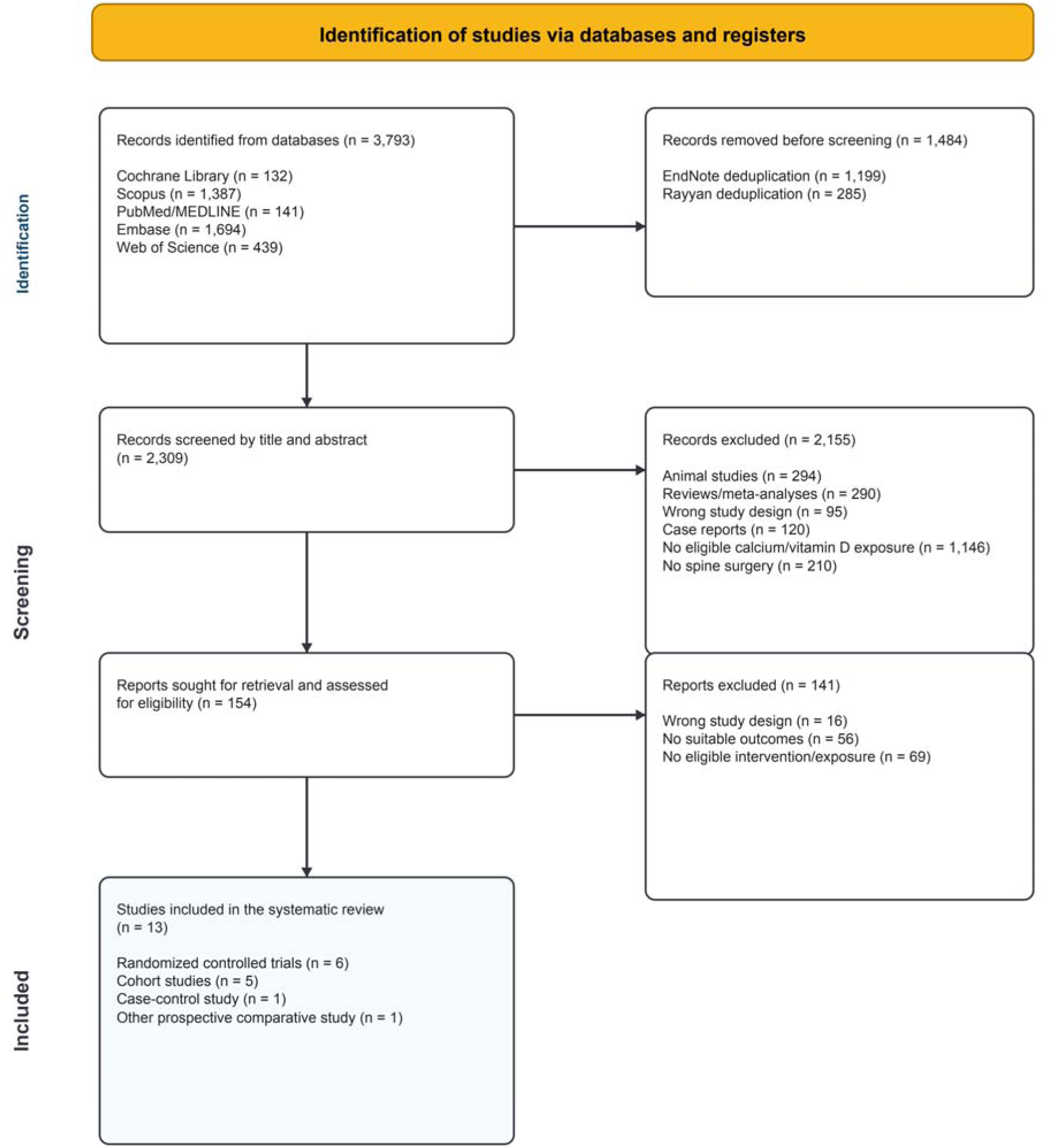
PRISMA 2020 flow diagram of study identification, screening, eligibility assessment, and inclusion. The diagram summarizes the number of records identified through database searching, duplicate records removed, records screened by title and abstract, reports assessed for full-text eligibility, reports excluded with reasons, and studies included in the systematic review and quantitative syntheses. PRISMA, Preferred Reporting Items for Systematic Reviews and Meta-Analyses.

### Characteristics of the included studies

The 13 included studies were published between 2014 and 2026 and represented a clinically and methodologically heterogeneous evidence base. The included reports comprised studies by Haddadi et al. [21], Hu et al. [22], Ko et al. [23], Krasowska et al. [24], Nekhlopochyn et al. [25], Shafiee et al. [26], and Shi et al. [27]. The remaining included studies were reported by Skrobot et al. [28,29], Xu et al. [30], Dzik et al. [31], Baumann et al. [32], and Li et al. [33].

Six studies were randomized controlled trials, five were prospective or retrospective cohort studies, one was a nonrandomized prospective comparative study, and one was a retrospective case-control study. The studies were conducted in Iran, Taiwan, South Korea, Poland, Ukraine, China, the United States, and Germany. Surgical populations included patients undergoing lumbar decompression, posterior lumbar fusion, posterior or transforaminal lumbar interbody fusion, anterior cervical interbody fusion, instrumented spinal stabilization, and lumbosacral fusion.

Vitamin D-related interventions varied substantially across studies. These included oral vitamin D alone, vitamin D combined with calcium, intramuscular vitamin D, activated vitamin D analogues, combined vitamin D and vitamin K treatment, different postoperative vitamin D doses, and vitamin D administered alongside rehabilitation or other anti-osteoporosis treatment. Comparator groups included placebo, calcium alone, routine care, no vitamin D correction, alendronate, different supplementation doses, and groups defined according to baseline vitamin D or bone status. Follow-up ranged from early postoperative assessment to two years or longer.

The principal outcomes included postoperative pain, disability, fusion rate, time to fusion, bone mineral density, implant loosening, cage migration or subsidence, pathological vertebral or sacral fracture, revision surgery, quality of life, inflammatory markers, muscle metabolism, balance, and postural stability. Characteristics of the included studies are summarized in Table 1.

**Table 1.**
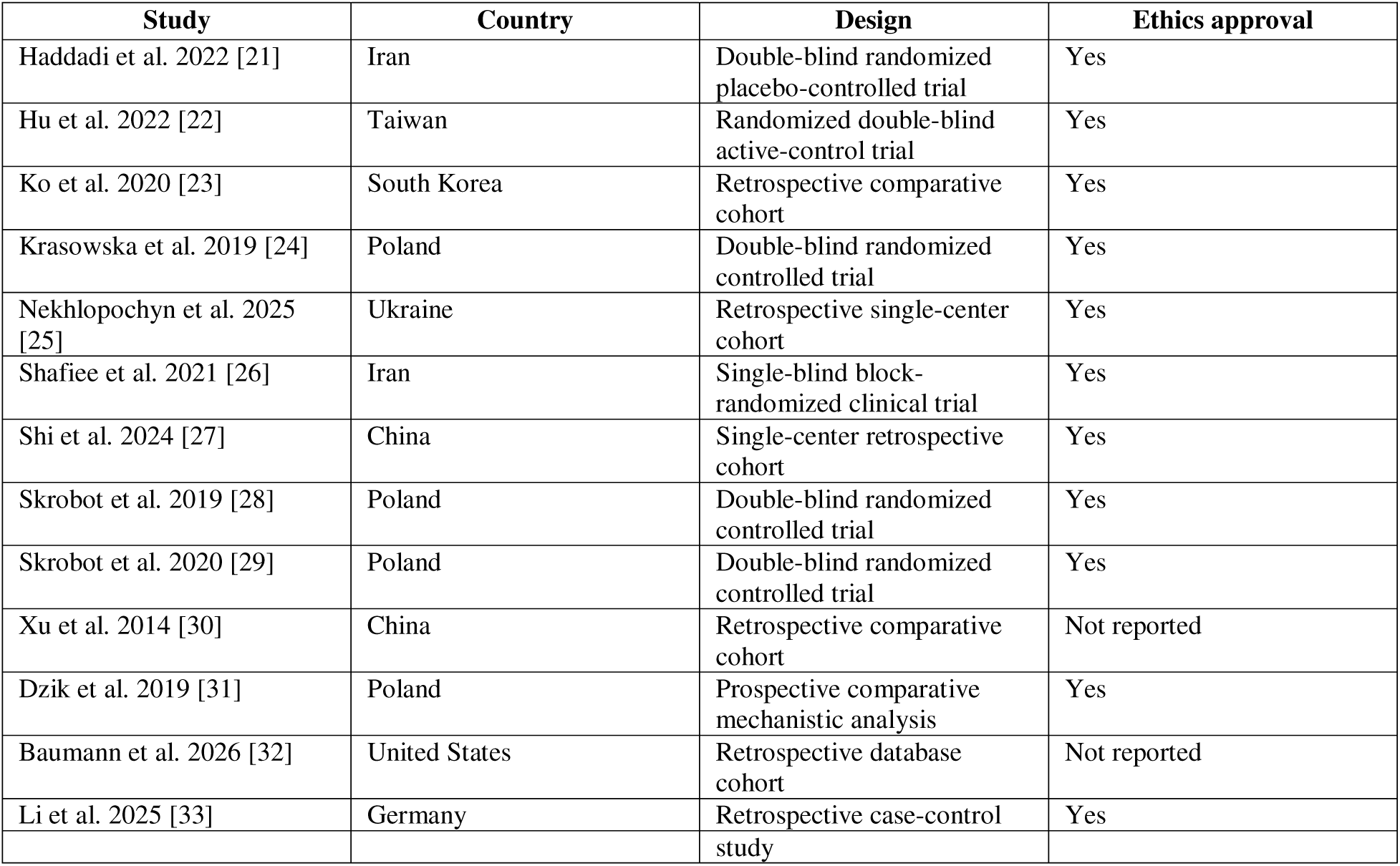
Characteristics of the 13 included studies.

#### Narrative clinical outcomes Fusion and time to fusion

Four included studies reported radiographic fusion outcomes. In the randomized trial by Hu et al., vitamin D combined with calcium citrate did not significantly increase the one-year fusion rate compared with calcium alone; however, the vitamin D group had a significantly shorter median time to fusion [22]. Shafiee et al. found no statistically significant difference in six-month nonfusion rates among patients receiving vitamin D, alendronate, or routine postoperative care [26].

In the cohort reported by Xu et al., postoperative treatment with 1,25-dihydroxyvitamin D was associated with higher fusion rates at six months and at final follow-up among osteoporotic patients undergoing transforaminal lumbar interbody fusion [30]. In the study by Shi et al., fusion outcomes differed between osteopenic patients receiving calcium and activated vitamin D and osteoporotic patients receiving the same background treatment together with denosumab [27]. Because this comparison involved differences in baseline bone status and denosumab co-treatment, the observed fusion difference could not be attributed independently to vitamin D.

### Implant-related complications and revision surgery

Nekhlopochyn et al. evaluated correction of vitamin D deficiency using a combined vitamin D, vitamin K, and omega-3 preparation in patients undergoing instrumented spinal stabilization [25]. Untreated vitamin D deficiency was associated with a greater burden of screw loosening, cage displacement, and revision procedures during follow-up. Patients with corrected deficiency had fewer progressive implant-related complications, although the multicomponent intervention and observational design limited attribution of the findings specifically to vitamin D.

Li et al. reported that patients who developed pathological vertebral fragility after lumbar fusion had received significantly lower daily vitamin D doses than controls [33]. Low-dose supplementation, defined as less than 2,000 IU/day, was associated with higher odds of pathological vertebral fragility than supplementation of at least 2,000 IU/day. Higher vitamin D doses were independently associated with a lower probability of fragility-related complications in multivariable analysis.

Revision surgery was reported most clearly by Nekhlopochyn et al. [25]. The untreated- deficiency group experienced increasing numbers of revision procedures between six and 12 months, together with screw loosening and cage displacement. Implant-related findings in the vitamin D-normal and treated-deficiency groups were described as less progressive and generally did not require surgical correction.

### Bone mineral density and vertebral bone quality

Bone-density and bone-quality findings were reported using bone mineral density measurements or computed-tomography-derived indices. In the cohort by Shi et al., bone mineral density decreased during six-month follow-up in the osteopenia group receiving calcium and activated vitamin D, whereas it increased in the osteoporosis group receiving the same background treatment together with denosumab [27]. This improvement could not be attributed to calcium or vitamin D alone because denosumab represented a major co-intervention.

Nekhlopochyn et al. reported an increase in computed-tomography-derived bone density among patients receiving vitamin D correction and a decrease among untreated comparators [25]. Li et al. found that vertebral Hounsfield-unit values were substantially lower among patients who developed pathological vertebral fragility than among controls [33]. Lower vitamin D□ doses were also associated with poorer vertebral bone quality, although age and other clinical differences between dose groups represented potential confounders.

### Pain, disability, and quality of life

Haddadi et al. compared vitamin D, alendronate, and routine care in women with sufficient baseline vitamin D concentrations undergoing lumbar fusion [21]. Visual Analog Scale and Oswestry Disability Index scores improved over time in all groups, but no statistically significant between-group differences were observed at six months.

Among vitamin D-deficient patients undergoing decompression for lumbar spinal stenosis, Ko et al. reported that intramuscular vitamin D□ was associated with lower Oswestry Disability Index scores and higher physical and mental component scores of the 36-Item Short Form Health Survey at 12 and 24 months [23]. Roland–Morris Disability Questionnaire scores did not differ significantly between groups.

Hu et al. reported lower Visual Analog Scale scores in the vitamin D-plus-calcium group at six months and lower Oswestry Disability Index scores at three and six months compared with calcium alone [22]. Shafiee et al. observed improvement in pain within all treatment groups, but differences in pain improvement between the vitamin D, alendronate, and routine-care groups were not statistically significant [26].

### Inflammatory and neuromuscular outcomes

Krasowska et al. evaluated five weeks of preoperative vitamin D supplementation before posterior lumbar interbody fusion [24]. Supplementation increased serum 25-hydroxyvitamin D concentrations. Following surgery and rehabilitation, reductions in C-reactive protein, interleukin-6, and tumor necrosis factor-α were reported in the supplemented group, whereas changes in interleukin-10 were not significant. Pain decreased after surgery and rehabilitation in both groups.

Two randomized trials by Skrobot et al. assessed balance and postural control after posterior lumbar interbody fusion and anterior cervical interbody fusion [28,29]. Vitamin D supplementation was associated with earlier or greater improvement in selected limits-of-stability and postural-stability measurements during postoperative rehabilitation. Effects on measured fall risk were inconsistent across the two studies.

Dzik et al. examined vitamin D status, preoperative supplementation, and multifidus muscle biomarkers in patients undergoing posterior lumbar interbody fusion [31]. Supplementation was associated with higher citrate synthase activity and changes in Akt and FOXO3a signaling consistent with reduced pro-atrophy signaling. Differences were also observed in insulin-like growth factor 1, atrogin-1, and markers of mitochondrial function. Radiographic fusion was not reported.

### Postoperative fracture outcomes

Baumann et al. evaluated 13,625 adults with osteoporosis undergoing first-time lumbosacral fusion [32]. Postoperative sacral fracture occurred in 347 patients, corresponding to a two-year incidence of 2.55%. Vitamin D prescription was associated with a lower risk of sacral fracture in adjusted proportional-hazards analysis (hazard ratio 0.775, 95% CI 0.603–0.997; p = 0.047). Bisphosphonate use was associated with increased fracture risk, while denosumab and teriparatide were not significantly associated with fracture occurrence. Increasing age and a history of falls were also independently associated with sacral fracture.

In the case-control study by Li et al., patients with pathological vertebral fragility received lower mean daily vitamin D doses than controls [33]. Low-dose supplementation was associated with higher odds of pathological vertebral fragility, particularly among older patients. Receiver-operating-characteristic analysis identified a vitamin D dose of approximately 1,900 IU/day as the threshold that best discriminated pathological vertebral fragility, although this observational threshold should not be interpreted as a definitive treatment recommendation.

### Safety

Safety reporting was limited and inconsistent. In the randomized trial by Hu et al., no vitamin D toxicity or hypercalcemia was reported [22]. Two postoperative infections occurred in the control group and resolved with antibiotic treatment. Xu et al. reported no fixation loosening or implant breakage during follow-up among osteoporotic patients treated with postoperative calcitriol [30]. Most other studies did not provide sufficiently detailed or standardized adverse-event reporting to permit comparative safety analysis.

A complete summary of the interventions, baseline vitamin D status, outcomes, and study-level findings is presented in Table 2.

**Table 2.**
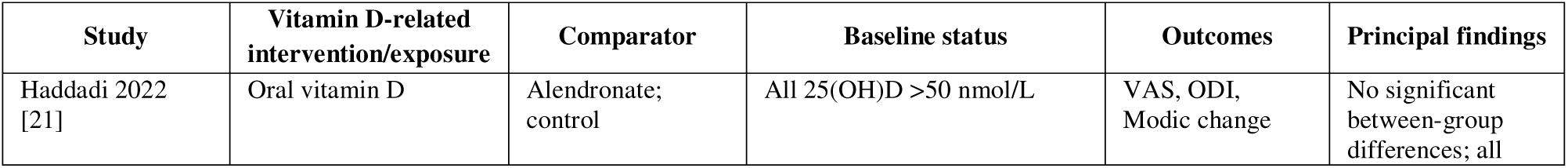
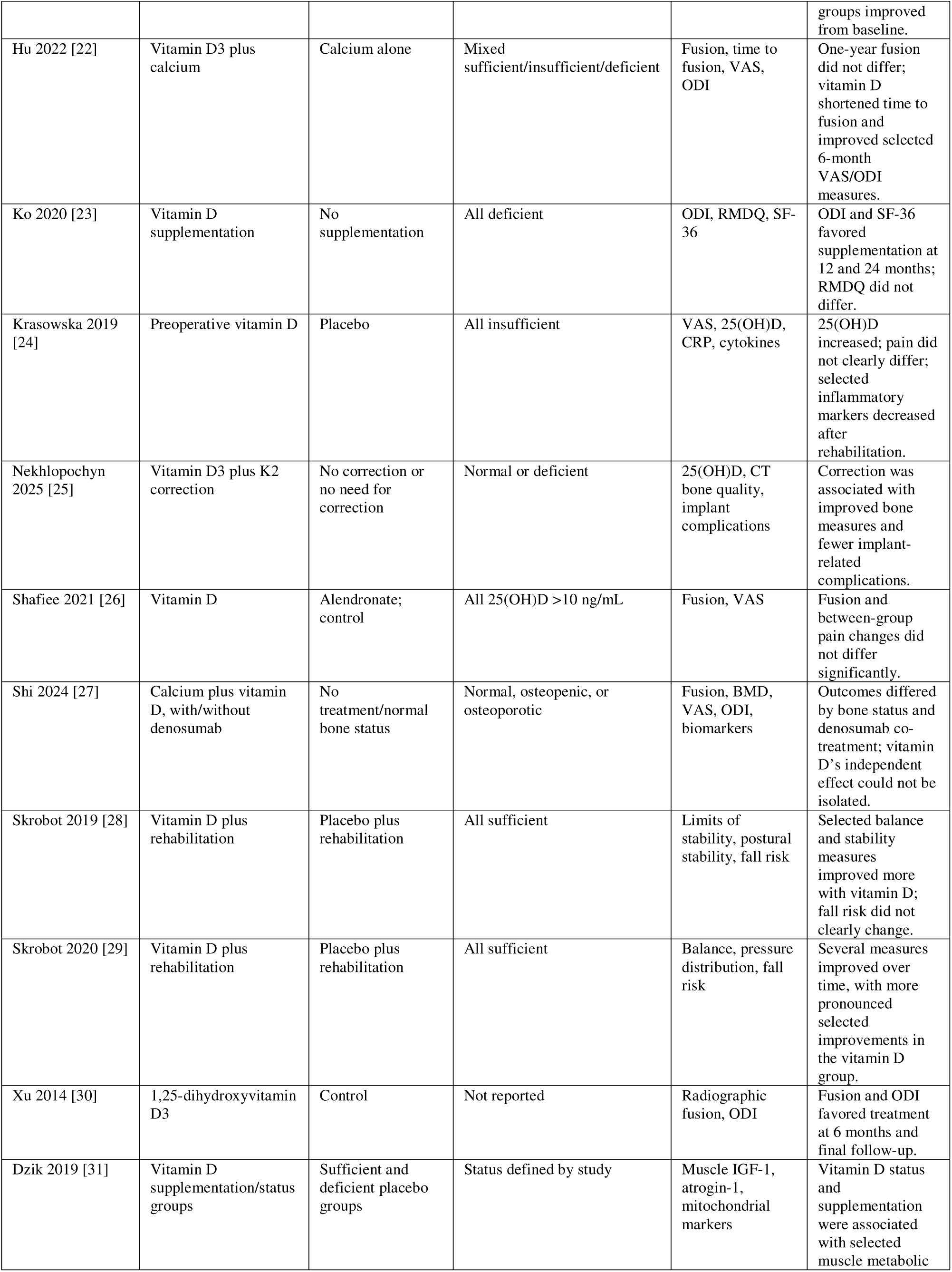
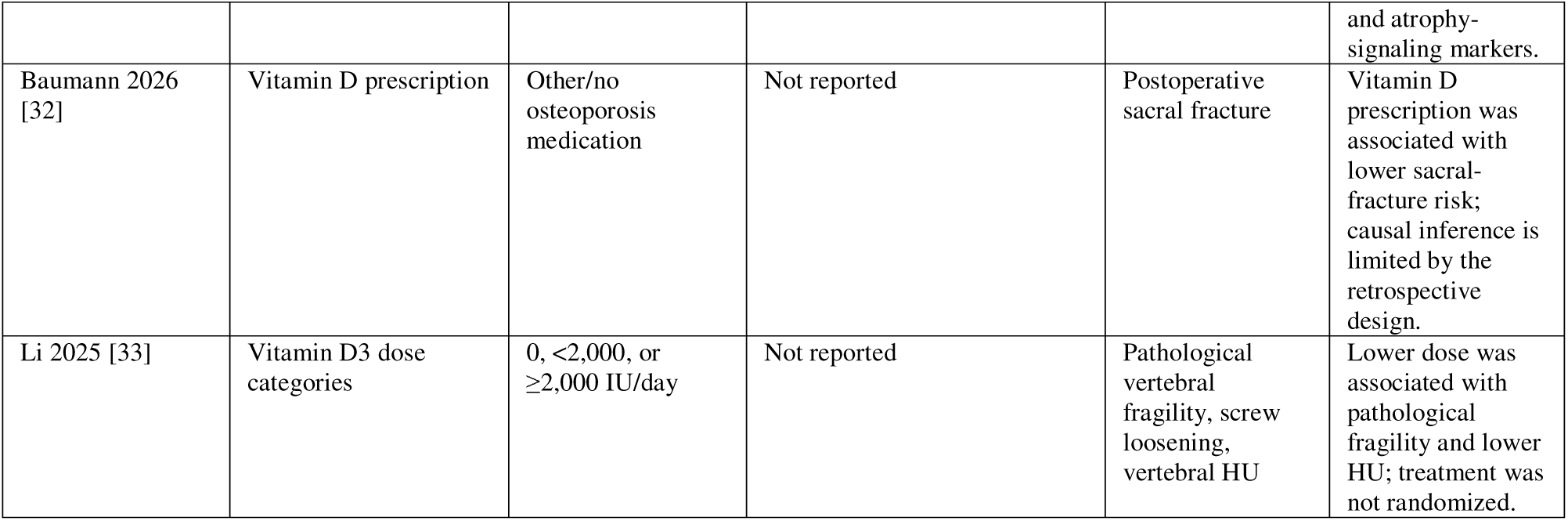
Vitamin D-related interventions or exposures, comparators, baseline vitamin D status, outcomes, and principal findings of the 13 included studies.

#### Primary quantitative outcome: postoperative pain

Four direct supplementation studies involving 177 participants contributed to the intervention-focused meta-analysis of postoperative pain measured using the Visual Analog Scale. The pooled random-effects estimate did not demonstrate a statistically significant difference between vitamin D supplementation and comparator groups (MD −0.82, 95% CI −2.50 to 0.86; p = 0.2176). Between-study heterogeneity was substantial (I² = 84.3%; τ = 0.9648), indicating marked variation in the magnitude and direction of the observed effects. The intervention-focused meta-analysis of postoperative Visual Analog Scale scores is presented in Figure 2.

**Figure 2.**
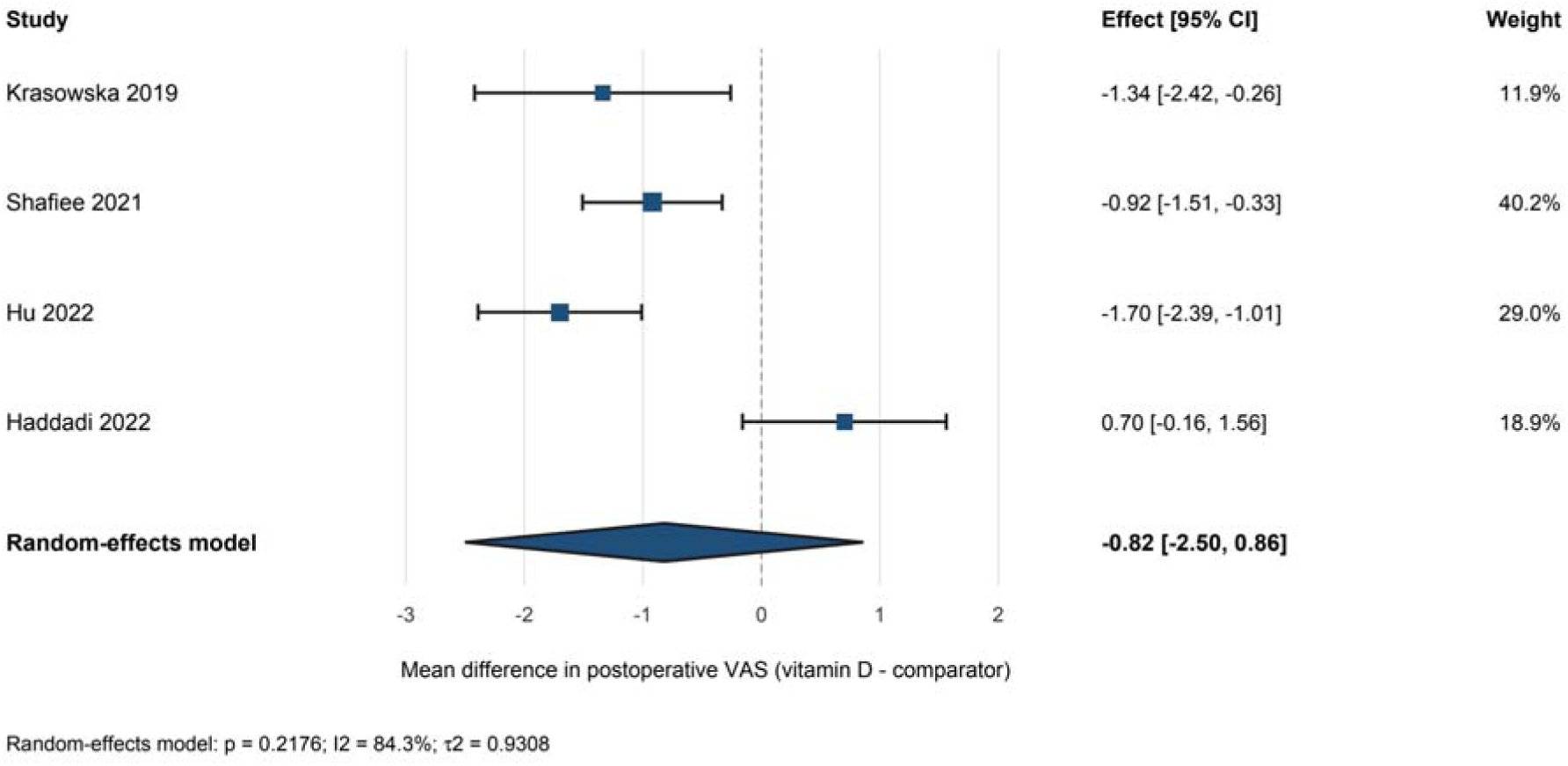
Forest plot of postoperative Visual Analog Scale scores in direct vitamin D supplementation studies. The figure presents study-specific and pooled mean differences in postoperative pain between vitamin D supplementation and comparator groups. Mean differences below zero favor vitamin D supplementation. Squares represent individual estimates, square size reflects study weight, horizontal lines represent 95% confidence intervals, and the diamond represents the random-effects estimate. CI, confidence interval; MD, mean difference; VAS, Visual Analog Scale.

An exploratory analysis incorporated the clinically distinct study by Shi et al., resulting in five studies and 342 participants. Four studies reported lower postoperative Visual Analog Scale scores in the vitamin D-related group, whereas Haddadi et al. reported an effect in the opposite direction. The pooled random-effects estimate remained statistically nonsignificant (MD −0.72, 95% CI −1.85 to 0.41; p = 0.1499). Heterogeneity remained substantial (I² = 84.2%; τ = 0.8124; Cochran Q p < 0.0001).

Influence diagnostics identified Haddadi et al. as the most influential study in the exploratory analysis. When this study was omitted, the pooled estimate favored the vitamin D-related group and reached statistical significance (MD −1.00, 95% CI −1.93 to −0.07; p = 0.0414). However, heterogeneity remained substantial, and the result was dependent on exclusion of a single study. This sensitivity finding was therefore considered hypothesis-generating rather than evidence of a robust analgesic effect.

The exploratory five-study analysis, leave-one-out analysis, influence diagnostics, and additional sensitivity analyses are provided in Supplementary Material 5.

### Secondary quantitative outcome: disability

Three studies involving 277 participants contributed to the exploratory meta-analysis of postoperative disability measured using the Oswestry Disability Index. These studies included direct supplementation and vitamin D or bone-status comparisons. The pooled random-effects model favored the vitamin D-related group (MD −6.40, 95% CI −10.41 to −2.39; p = 0.0017). The exploratory meta-analysis of postoperative Oswestry Disability Index scores is presented in Figure 3.

**Figure 3.**
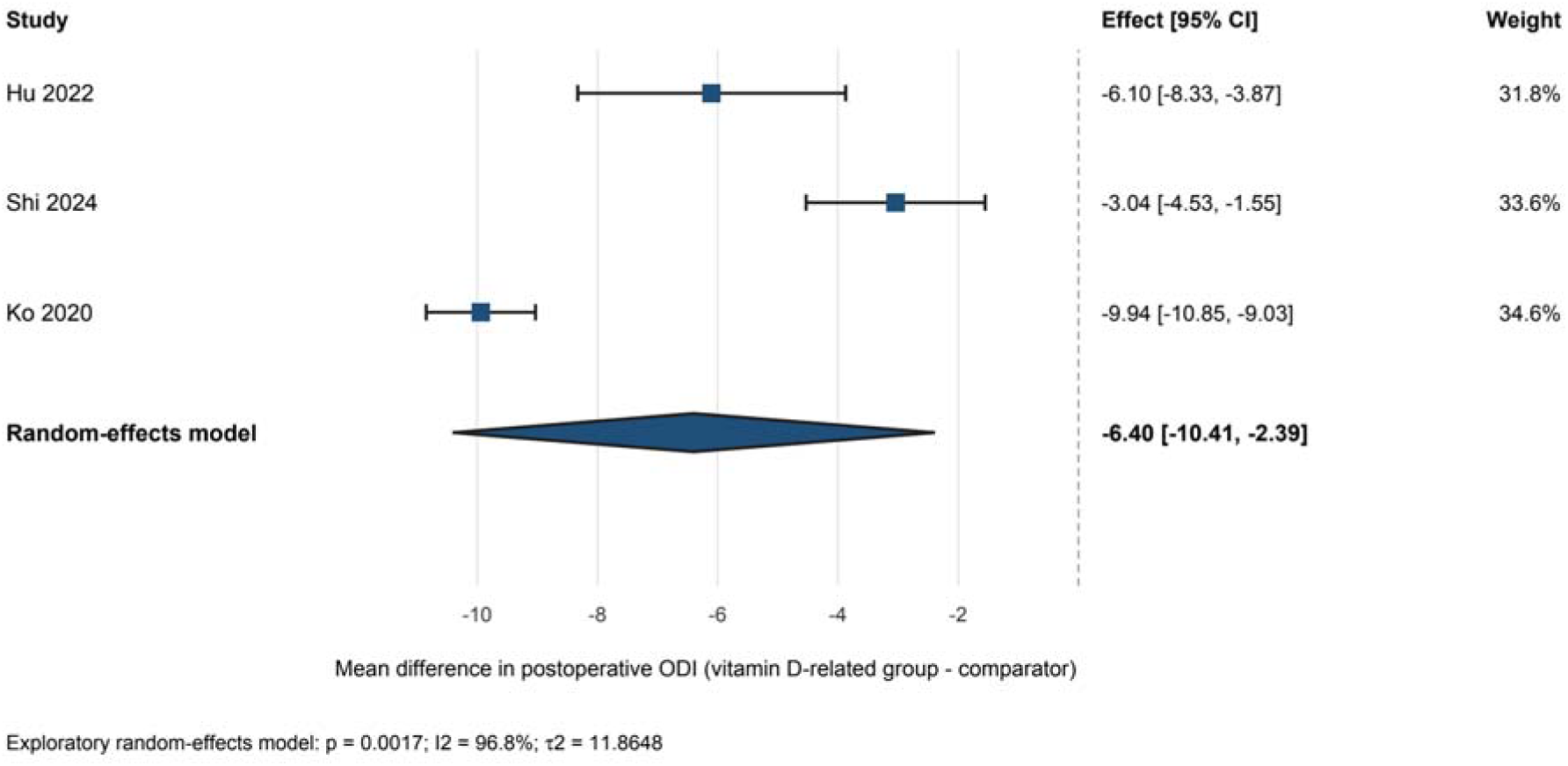
Forest plot of postoperative Oswestry Disability Index scores. The figure presents study-specific and pooled mean differences in postoperative disability between vitamin D-related or more favorable vitamin D or bone-metabolic groups and their comparators. Mean differences below zero favor the vitamin D-related or more favorable-status group. Because the studies combined direct supplementation with vitamin D or bone-status comparisons, the analysis was considered exploratory. CI, confidence interval; MD, mean difference; ODI, Oswestry Disability Index.

Between-study heterogeneity was very high (I² = 96.8%; τ² = 11.8648), indicating substantial variability in study design, population, exposure definition, comparator selection, surgical procedure, and timing of outcome measurement. The pooled estimate should therefore not be interpreted as a uniform treatment effect.

Leave-one-out analysis showed that the direction of association remained favorable after sequential omission of individual studies. However, statistical significance was partially dependent on Hu et al. When Hu et al. was omitted, the pooled estimate was attenuated and the confidence interval crossed the null (MD −6.52, 95% CI −13.28 to 0.25; p = 0.0589). Omission of Shi et al. strengthened the pooled effect (MD −8.16, 95% CI −11.91 to −4.41; p < 0.0001), while omission of Ko et al. produced an estimate of MD −4.45 (95% CI −7.44 to −1.46; p = 0.0035).

These findings suggest a possible association between vitamin D-related exposure and lower postoperative disability, but the very high heterogeneity and sensitivity to individual-study omission substantially limit confidence in the pooled estimate. The leave-one-out sensitivity analysis and exploratory funnel plot are provided in Supplementary Material 5.

### Secondary quantitative outcome: fusion rate

Three direct supplementation or treatment studies involving 128 participants contributed to the intervention-focused fusion-rate analysis. The pooled random-effects estimate favored vitamin D-containing treatment but was not statistically significant (RR 1.17, 95% CI 0.78–1.74; p = 0.2436). Heterogeneity was low to moderate (I² = 31.5%; τ² = 0.0062; Cochran Q p = 0.2322). The intervention-focused meta-analysis of fusion rate is presented in Figure 4.

**Figure 4.**
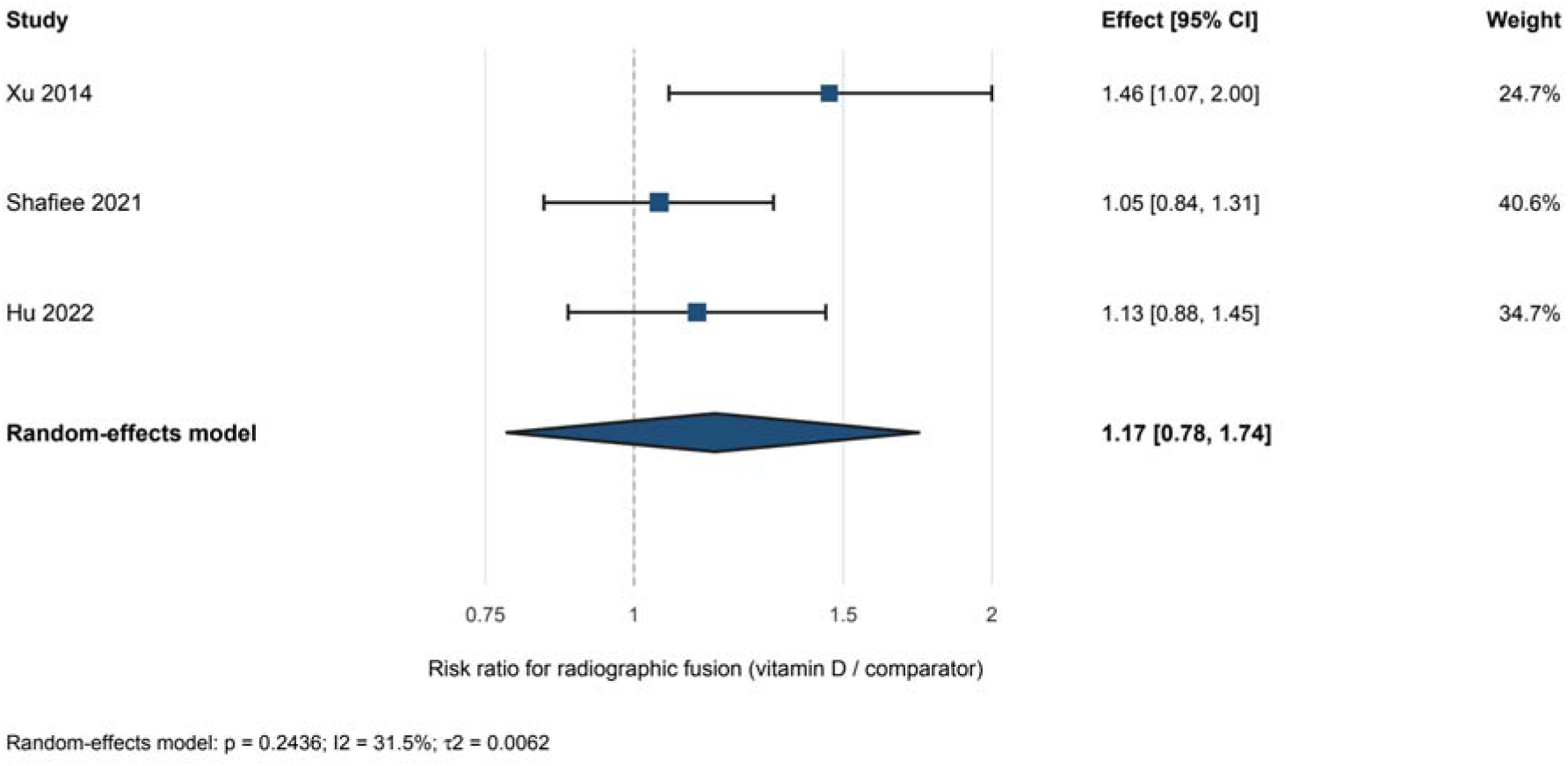
Forest plot of radiographic fusion in direct vitamin D supplementation or treatment studies. The figure presents study-specific and pooled risk ratios for radiographic fusion. Risk ratios greater than 1 favor vitamin D-containing treatment. Squares represent individual estimates, square size reflects study weight, horizontal lines represent 95% confidence intervals, and the diamond represents the random-effects estimate. CI, confidence interval; RR, risk ratio.

An exploratory analysis additionally incorporated the clinically heterogeneous bone-status and co-intervention comparison reported by Shi et al. Four studies involving 293 participants contributed to this analysis, including 119 participants in the vitamin D-related or more favorable-status groups and 174 in the comparator groups. The pooled estimate favored the vitamin D-related groups but did not reach statistical significance (RR 1.24, 95% CI 0.98–1.57; p = 0.0617). Moderate heterogeneity was observed (I² = 45.5%; τ² = 0.0099; Cochran Q p = 0.1385), and the prediction interval ranged from 0.83 to 1.84.

The comparison reported by Shi et al. produced an RR of 1.36 (95% CI 1.19–1.55). However, this estimate was derived from a single study in which the groups differed in baseline bone status and denosumab exposure. It should therefore be interpreted descriptively rather than as an isolated vitamin D effect. The test for subgroup differences between supplementation studies and the clinically distinct bone-status comparison was not statistically significant (p = 0.1836), although the analysis was underpowered.

Leave-one-out analysis of the four-study dataset yielded pooled estimates ranging from RR 1.17 to RR 1.32. The direction of association remained above 1 after omission of each individual study, but statistical significance was sensitive to study exclusion. Omission of Shafiee et al. produced an RR of 1.32 (95% CI 1.03–1.69) and reduced heterogeneity to 6.3%. Omission of Shi et al. returned the estimate to that of the supplementation-only analysis (RR 1.17, 95% CI 0.78– 1.74; I² = 31.5%).

Within the supplementation-only analysis, leave-one-out pooled estimates ranged from RR 1.08 to RR 1.26. All confidence intervals crossed the null and were wide because only two studies remained in each omission scenario. Omission of Xu et al. reduced detected heterogeneity to 0% and produced an RR of 1.08 (95% CI 0.69–1.70). Although no statistical heterogeneity was detected in this scenario, the pooled estimate remained imprecise and statistically nonsignificant.

A sensitivity analysis using the six-month fusion data reported by Xu et al. produced the same pooled point estimate as the exploratory analysis (RR 1.24), with a wider confidence interval (95% CI 0.93–1.63). Heterogeneity increased to 51.5%, and the prediction interval ranged from 0.79 to 1.92. The selected follow-up time therefore did not materially alter the direction or magnitude of the pooled estimate, but considerable uncertainty remained.

The exploratory combined analysis, subgroup analysis, leave-one-out analyses, and six-month sensitivity analysis are provided in Supplementary Material 5.

### Small-study effects

Funnel plots were generated for the Visual Analog Scale, Oswestry Disability Index, exploratory overall fusion-rate, and supplementation-only fusion-rate analyses. However, only three to five studies contributed to each outcome. With fewer than 10 studies, visual interpretation of funnel plots and trim-and-fill analyses has limited reliability and may reflect chance, clinical heterogeneity, or differences in study precision rather than publication bias.

Accordingly, no firm conclusion regarding publication bias or small-study effects was drawn. The exploratory funnel plots and trim-and-fill analysis are provided in Supplementary Material 5.

### Risk-of-bias assessment

Risk of bias was assessed using the Joanna Briggs Institute critical appraisal checklist appropriate to each study design. Six randomized controlled trials, five cohort studies, one nonrandomized prospective comparative study, and one case-control study were assessed.

Two studies were categorized as having low risk of bias, eight as having moderate risk, and three as having high risk. Common methodological concerns included unclear allocation concealment, incomplete blinding, limited control of confounding, small sample sizes, incomplete follow-up reporting, imprecise exposure classification, and inconsistent outcome assessment. Observational studies were particularly limited by treatment-selection bias and residual confounding, while several randomized trials provided insufficient information regarding randomization, allocation concealment, or outcome-assessor blinding.

Detailed study-level risk-of-bias findings are presented in Table 3, and the complete item-level Joanna Briggs Institute assessments are provided in Supplementary Material 4.

**Table 3.**
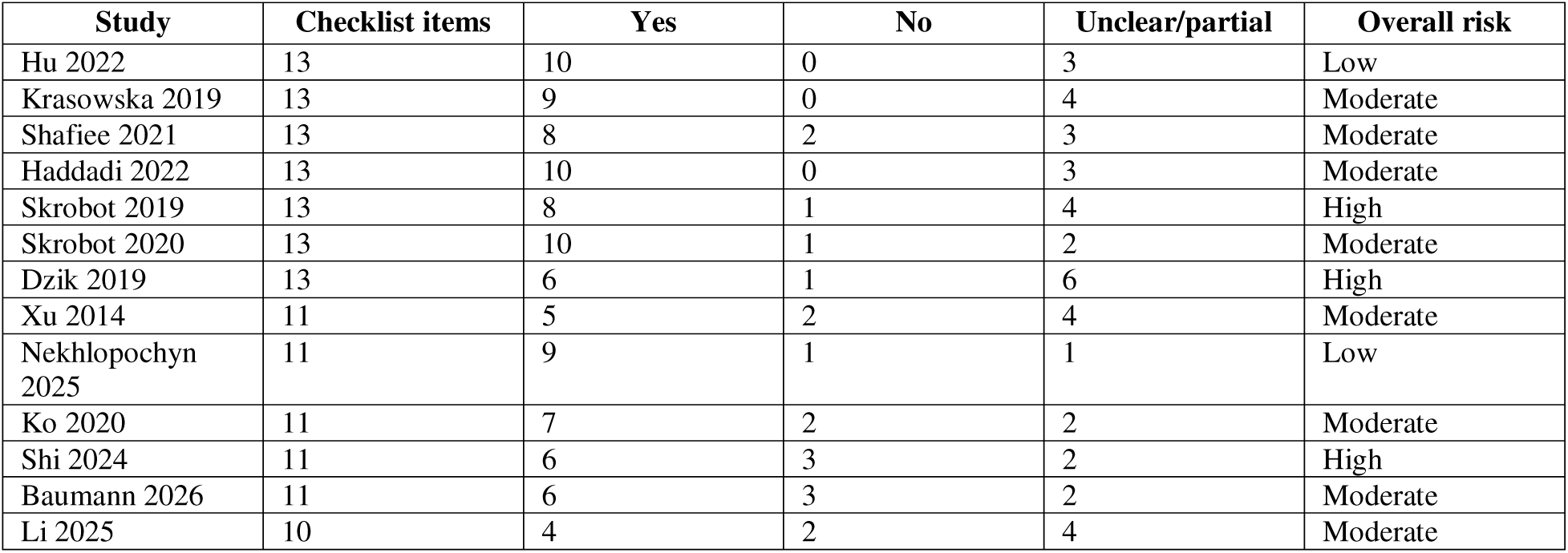
Joanna Briggs Institute risk-of-bias summary. Item-level judgments are provided in Supplementary Material 4. JBI, Joanna Briggs Institute.

## Discussion

### Principal findings

This systematic review and meta-analysis evaluated the association of vitamin D status and vitamin D-containing supplementation with clinical, functional, and bone-related outcomes after spine surgery. The principal quantitative finding was that direct vitamin D supplementation was not associated with a statistically significant reduction in postoperative pain. In the intervention-focused analysis of four studies involving 177 participants, the pooled mean difference in Visual Analog Scale scores was −0.82 points, but the confidence interval was wide and crossed the null. Similarly, the intervention-focused fusion analysis of three studies involving 128 participants did not demonstrate a statistically significant improvement in fusion rate.

An exploratory analysis of postoperative disability showed lower Oswestry Disability Index scores among patients receiving vitamin D supplementation or having more favorable vitamin D or bone-metabolic status. However, this finding was accompanied by very high heterogeneity and was sensitive to the omission of an individual study. The broader exploratory fusion analysis also favored vitamin D-related exposure but did not reach statistical significance and incorporated a clinically distinct comparison involving differences in baseline bone status and denosumab treatment.

Beyond the pooled outcomes, individual studies reported potentially favorable associations with shorter time to fusion, improved bone quality, fewer implant-related complications, lower postoperative fracture risk, and selected neuromuscular or rehabilitation outcomes. These findings suggest that vitamin D may influence several aspects of postoperative recovery, but the current evidence does not establish that supplementation independently improves pain or fusion success.

### Postoperative pain and disability

Postoperative pain was designated as the primary quantitative outcome. Although several individual studies reported lower Visual Analog Scale scores among patients receiving vitamin D-related treatment, the intervention-focused random-effects analysis did not demonstrate a statistically significant overall effect. Considerable heterogeneity was present, reflecting differences in baseline vitamin D status, supplementation regimen, comparator group, surgical procedure, follow-up duration, and timing of postoperative pain assessment.

Haddadi et al. evaluated vitamin D, alendronate, and routine care in patients with sufficient baseline vitamin D concentrations and found improvement over time in all groups without a significant between-group difference [21]. In contrast, Hu et al. reported lower six-month pain scores among patients receiving vitamin D together with calcium than among those receiving calcium alone [22]. Ko et al. primarily evaluated disability and quality of life in vitamin D-deficient patients undergoing decompression for lumbar spinal stenosis, whereas Krasowska et al. examined perioperative pain together with inflammatory responses following preoperative vitamin D supplementation [23,24]. These studies addressed different populations and clinical questions, limiting interpretation of a single pooled analgesic effect.

In the broader five-study exploratory analysis, the pooled estimate remained nonsignificant. Influence diagnostics identified Haddadi et al. as the most influential study, and omission of this study resulted in a statistically significant pooled difference. However, statistical significance after removal of one influential study should not be interpreted as confirmation of treatment efficacy. Haddadi et al. evaluated a clinically relevant population with sufficient baseline vitamin D concentrations, and its findings may indicate that supplementation provides limited additional analgesic benefit in patients who are not deficient. The sensitivity result is therefore more appropriately interpreted as suggesting that baseline vitamin D status may modify the observed association rather than as justification for excluding the study from the primary interpretation.

The exploratory Oswestry Disability Index analysis produced a pooled mean difference of −6.40 points in favor of vitamin D-related exposure. This magnitude may be relevant to postoperative functional recovery, although its clinical importance depends on baseline disability, follow-up duration, and the threshold used to define a meaningful individual change. The analysis had very high heterogeneity, and omission of Hu et al. caused the confidence interval to cross the null. The pooled finding should therefore be viewed as a possible association rather than definitive evidence that supplementation improves disability.

Disability after spine surgery is influenced by pain, neural decompression, muscle function, psychological status, rehabilitation, surgical complications, and baseline disease severity. Vitamin D supplementation may contribute to recovery in deficient patients through skeletal and neuromuscular pathways, but the available studies do not permit its independent effect to be reliably separated from these factors.

### Fusion and bone-healing outcomes

The intervention-focused fusion analysis did not demonstrate a statistically significant improvement in fusion rate. The pooled risk ratio of 1.17 favored vitamin D-containing treatment, but the confidence interval was wide and compatible with both no meaningful effect and a potentially important benefit. The small number of participants and fusion events limited the precision of the estimate.

Individual fusion studies produced inconsistent findings. Hu et al. reported that vitamin D combined with calcium did not significantly improve the final one-year fusion rate compared with calcium alone, although the vitamin D group achieved fusion earlier [22]. Median time to fusion was approximately 169 days in the vitamin D group and 185 days in the control group. Earlier fusion may be clinically relevant even when the final proportion achieving fusion is similar, because prolonged incomplete arthrodesis may extend activity restrictions and expose the instrumentation to sustained mechanical stress. Nevertheless, whether a difference of this magnitude translates into fewer complications or better long-term outcomes remains uncertain.

Shafiee et al. found no significant difference in six-month nonfusion rates among patients receiving vitamin D, alendronate, or routine care [26]. The inclusion of patients without clearly characterized severe deficiency may have reduced the likelihood of observing a supplementation effect. By contrast, Xu et al. reported higher fusion rates at six months and final follow-up among osteoporotic patients receiving postoperative 1,25-dihydroxyvitamin D [30]. This study supports a possible benefit in a metabolically vulnerable population, but its observational design and limited sample size reduce certainty regarding causality.

Shi et al. reported better fusion outcomes in osteoporotic patients receiving calcium, activated vitamin D, and denosumab than in osteopenic patients receiving calcium and activated vitamin D alone [27]. This comparison was included only in the exploratory synthesis because the groups differed in baseline bone status and the osteoporosis group received denosumab. Denosumab can substantially influence bone turnover and bone mineral density, preventing the observed difference from being attributed independently to vitamin D.

The four-study exploratory fusion estimate was stronger than the supplementation-only estimate but remained statistically nonsignificant. Its prediction interval crossed the null, indicating that a future comparable study could plausibly demonstrate no benefit, a modest adverse association, or a clinically important favorable association. The stronger estimate was partly driven by the clinically distinct study by Shi et al., emphasizing the importance of separating direct supplementation effects from associations involving baseline bone status or major co-interventions.

Leave-one-out analyses showed that the direction of the exploratory fusion estimate remained above 1 after sequential omission of each study. However, statistical significance was achieved only after omission of Shafiee et al., and the supplementation-only estimates remained imprecise in all omission scenarios. These findings demonstrate directional consistency but not statistical robustness.

### Bone quality, implant stability, and fracture outcomes

Several observational studies suggested that vitamin D-related exposure may be associated with bone quality and mechanical outcomes beyond the final fusion rate. Nekhlopochyn et al. reported improved computed-tomography-derived bone density and fewer implant-related complications among patients receiving correction of vitamin D deficiency with a combined vitamin D□, vitamin K□, and omega-3 preparation [25]. Untreated deficiency was associated with more screw loosening, cage displacement, and revision procedures. However, the intervention contained several biologically active components, and treatment acceptance was not randomized. Differences in adherence, nutritional status, comorbidity, and clinical management may therefore have influenced the observed associations.

Baumann et al. evaluated a large multicenter cohort of adults with osteoporosis undergoing lumbosacral fusion [32]. Vitamin D prescription was associated with a lower adjusted risk of postoperative sacral fracture, whereas increasing age and a history of falls were associated with higher risk. The large sample size strengthens the precision of the association, but prescription data do not confirm adherence, achieved serum vitamin D concentrations, or correction of deficiency. Confounding by health behavior and treatment selection also remains possible.

Li et al. found that patients who developed pathological vertebral fragility had received lower postoperative vitamin D□ doses and had lower vertebral Hounsfield-unit measurements than controls [33]. Higher daily doses were associated with lower fragility risk, and an exploratory threshold of approximately 1,900 IU/day was identified. Because this threshold was derived retrospectively from a single cohort, it should not be interpreted as an established optimal dose. Patients receiving different doses also differed in age and bone quality, and residual confounding cannot be excluded.

Collectively, these observational findings suggest that vitamin D status or treatment may be associated with bone–implant integration, vertebral strength, and fracture risk. However, they do not establish that increasing the supplementation dose directly prevents mechanical complications. Randomized dose-comparison studies incorporating serum monitoring and standardized radiographic outcomes are required.

### Inflammatory, muscular, and rehabilitation-related outcomes

The clinical relevance of vitamin D after spine surgery may extend beyond bone mineralization. Krasowska et al. reported that preoperative supplementation increased serum 25-hydroxyvitamin D concentrations and was followed by reductions in selected inflammatory markers during postoperative rehabilitation [24]. These changes were not accompanied by a clearly established independent analgesic effect, but they suggest a possible interaction between vitamin D status and perioperative inflammatory recovery.

Two randomized studies by Skrobot et al. evaluated balance and postural control during rehabilitation after lumbar and cervical interbody fusion [28,29]. Vitamin D supplementation was associated with earlier or greater improvement in selected balance measures, although findings for fall-risk outcomes were inconsistent. These studies were small and assessed multiple outcomes, increasing the possibility of chance findings. Nevertheless, the observed changes are biologically compatible with the known role of vitamin D in muscle function and neuromuscular coordination.

Dzik et al. examined vitamin D status and multifidus muscle biology in patients undergoing posterior lumbar interbody fusion [31]. Supplementation was associated with differences in citrate synthase activity and Akt–FOXO3a signaling consistent with improved mitochondrial activity and reduced pro-atrophy signaling. These mechanistic findings support a possible biological pathway through which vitamin D could influence postoperative muscle recovery. However, muscle biomarkers are surrogate outcomes, and radiographic fusion or major clinical endpoints were not evaluated.

### Biological interpretation

Vitamin D has several biological functions relevant to recovery after spine surgery. Through the vitamin D receptor, its active metabolite, 1,25-dihydroxyvitamin D, influences intestinal calcium absorption, parathyroid hormone regulation, osteoblast differentiation, osteoclast signaling, mineralization, and skeletal remodeling [34]. Spinal arthrodesis depends on vascularization, osteoconduction, osteoinduction, new bone formation, and remodeling across the fusion bed. Severe vitamin D deficiency could theoretically impair one or more of these processes.

Vitamin D may also affect skeletal muscle strength, balance, proprioception, and inflammatory signaling. These effects could influence rehabilitation, fall risk, functional recovery, and mechanical loading of spinal instrumentation. The potential clinical effect of supplementation is therefore unlikely to be limited to the binary presence or absence of radiographic fusion.

However, biological plausibility does not establish clinical efficacy. The effect of supplementation is likely to depend on baseline deficiency, osteoporosis severity, supplementation dose and duration, adherence, calcium intake, renal function, concurrent anti-osteoporosis therapy, and treatment timing relative to surgery. Supplementation may provide little additional benefit in patients who are already vitamin D sufficient, whereas correction of severe deficiency may be more clinically relevant. The available aggregate data were insufficient to identify these potential treatment-effect modifiers.

### Comparison with previous evidence

Evidence from nonsurgical populations has suggested that vitamin D supplementation may improve selected measures of muscle strength and balance, although effects vary according to the population and baseline vitamin D status. A previous systematic review by Muir and Montero-Odasso reported potential benefits for muscle function and balance among older adults but did not establish uniform reductions in falls across all populations [35]. The small postoperative rehabilitation trials included in the present review are broadly compatible with a possible neuromuscular effect but remain insufficient to determine whether supplementation prevents falls after spine surgery.

The findings also align with the systematic review by Khalooeifard et al., which evaluated vitamin D deficiency among patients undergoing elective spinal fusion [36]. That review identified associations between deficiency and poorer functional outcomes but emphasized heterogeneity and insufficient evidence regarding supplementation, fusion, and pseudarthrosis. The present analysis extends that evidence by separating intervention-focused estimates from exploratory analyses combining supplementation, baseline status, and co-intervention comparisons. This distinction demonstrates that the apparent association is weaker and less certain when the analysis is restricted to direct supplementation studies.

### Clinical implications

The current evidence does not support routine vitamin D supplementation as a proven method for increasing fusion rates or reducing postoperative pain in all patients undergoing spine surgery. In particular, the available data do not demonstrate a clear benefit among patients with sufficient baseline vitamin D concentrations.

Nevertheless, identifying and correcting clinically important vitamin D deficiency remains reasonable as part of comprehensive perioperative bone-health optimization, especially in patients with osteoporosis, fragility fractures, poor nutritional status, or planned multilevel instrumented fusion. Correction of deficiency is biologically justified and may provide broader skeletal and neuromuscular benefits, but it should not replace established evaluation and treatment of osteoporosis.

Clinical decisions should account for baseline serum 25-hydroxyvitamin D concentration, calcium status, renal function, bone mineral density, fracture history, and concurrent bone-modifying treatment. Vitamin D and calcium should be considered components of a broader strategy that may also include nutritional optimization, smoking cessation, exercise, fall prevention, and appropriate antiresorptive or anabolic treatment.

The findings do not establish a specific vitamin D dose, target serum concentration, route, or treatment duration for patients undergoing spine surgery. Dose thresholds derived from retrospective studies should not be adopted as clinical recommendations without prospective validation.

### Future directions

Future trials should enroll sufficiently large and clinically well-characterized populations and stratify randomization according to baseline serum 25-hydroxyvitamin D concentration and bone mineral density. Patients with severe deficiency, insufficiency, and sufficient vitamin D concentrations should be analyzed separately because the effects of supplementation may differ substantially among these groups.

Interventions should use standardized vitamin D formulations, doses, initiation times, and treatment durations. Adherence and achieved serum vitamin D concentrations should be reported. Calcium intake and concurrent treatment with bisphosphonates, denosumab, teriparatide, or other bone-modifying agents should be standardized or incorporated into the analytical design.

Fusion should be assessed at predefined intervals using standardized computed-tomography or validated radiographic criteria, with blinded outcome assessment whenever possible. Studies should report time to fusion in addition to final fusion rate because supplementation may influence the rate of bone healing without changing the eventual proportion of patients achieving fusion.

Future studies should also evaluate implant loosening, cage subsidence, proximal junctional failure, vertebral fracture, revision surgery, pain, disability, balance, quality of life, adverse events, and cost-effectiveness. An individual participant data meta-analysis may ultimately be required to determine whether baseline deficiency, osteoporosis severity, age, surgical procedure, supplementation dose, or concurrent treatment modifies the association between vitamin D and postoperative outcomes.

### Limitations

This review has several limitations. First, only 13 studies met the eligibility criteria, and few studies contributed to each quantitative outcome. The intervention-focused pain analysis included four studies, the disability analysis included three studies, and the intervention-focused fusion analysis included three studies. The limited evidence base reduced statistical power and produced imprecise confidence intervals.

Second, clinical and methodological heterogeneity was substantial. Studies differed in surgical indication, spinal region, operative procedure, baseline vitamin D status, osteoporosis severity, supplementation formulation, dose, route, timing, treatment duration, comparator group, follow-up duration, and outcome definition. The pain and disability analyses had particularly high heterogeneity, limiting interpretation of the pooled estimates as common treatment effects.

Third, studies evaluating direct supplementation were clinically different from those evaluating baseline vitamin D status, supplementation dose, or broader bone-metabolic status. These comparisons estimate related but non-equivalent effects. The review addressed this limitation by prioritizing intervention-focused analyses and classifying combined analyses as exploratory, but some outcome syntheses remained heterogeneous.

Fourth, major co-interventions limited attribution of several findings. The study by Shi et al. included denosumab in the osteoporosis group, whereas the treatment evaluated by Nekhlopochyn et al. combined vitamin D□ with vitamin K and omega-3. Consequently, favorable outcomes in these studies cannot be attributed independently to vitamin D.

Fifth, several studies were observational and susceptible to confounding by indication, treatment-selection bias, adherence differences, nutritional status, age, baseline bone quality, comorbidity, and postoperative management. Even in adjusted analyses, residual and unmeasured confounding may have influenced the reported associations.

Sixth, methodological quality ranged from low to high risk of bias. Common concerns included unclear allocation concealment, incomplete blinding, small sample sizes, incomplete control of confounding, imprecise exposure classification, and inconsistent outcome assessment. Two studies were categorized as having low risk of bias, eight as having moderate risk, and three as having high risk.

Seventh, sensitivity analyses demonstrated partial dependence on individual studies. Exclusion of Haddadi et al. changed the statistical significance of the exploratory pain estimate, omission of Hu et al. attenuated the disability result, and omission of Shafiee et al. changed the statistical significance of the exploratory fusion analysis. These findings indicate that the pooled results were not fully robust.

Eighth, publication bias could not be assessed reliably. Each outcome included substantially fewer than 10 studies, making funnel-plot interpretation and trim-and-fill analysis unreliable. Apparent asymmetry may have reflected clinical heterogeneity, chance, or differences in study precision rather than selective publication.

Ninth, fusion definitions and assessment methods were not standardized. Studies used plain radiography, computed tomography, combined imaging, and different grading systems at varying follow-up intervals. These differences may have influenced event classification and contributed to heterogeneity.

Finally, aggregate study-level data did not permit adequate adjustment for baseline vitamin D concentration, osteoporosis severity, age, smoking, nutrition, renal function, surgical complexity, number of fused levels, instrumentation strategy, concurrent medication, or adherence. The findings should therefore be interpreted as associations derived from limited and heterogeneous evidence rather than definitive causal treatment effects.

## Conclusion

Current evidence does not establish that vitamin D supplementation independently reduces postoperative pain or improves fusion rates in patients undergoing spine surgery. Although exploratory analyses suggested possible associations with lower disability, shorter time to fusion, improved bone quality, fewer implant-related complications, and reduced postoperative fracture risk, these findings were derived from small and clinically heterogeneous studies and were frequently influenced by baseline vitamin D status, concurrent treatment, and residual confounding.

Assessment and correction of clinically important vitamin D deficiency may remain reasonable components of comprehensive perioperative bone-health optimization, particularly in patients with osteoporosis or other risk factors for impaired skeletal healing. However, the available evidence is insufficient to define an optimal supplementation regimen or support routine supplementation as a stand-alone strategy for improving spinal fusion outcomes. Adequately powered randomized trials stratified by baseline vitamin D status and using standardized supplementation protocols, fusion definitions, follow-up intervals, and patient-centered outcomes are required.

## Statements and Declarations

## Data Availability

All aggregate data supporting the findings of this systematic review and meta-analysis are included in the article and its supplementary materials. Supplementary Material 1 contains the complete database search strategies. Supplementary Material 2 contains the finalized decisions for all 154 full-text reports, including the 13 included studies and 141 exclusions grouped into the three PRISMA categories. Supplementary Material 3 contains the complete structured study-level data-extraction dataset in a single worksheet. Supplementary Material 4 contains the complete item-level Joanna Briggs Institute risk-of-bias assessments. Supplementary Material 5 contains the supplementary forest plots, subgroup and sensitivity analyses, influence diagnostics, and small-study-effect plots. Supplementary Material 6 contains the completed PRISMA 2020 checklist, indicating where each reporting item is addressed in the manuscript or supplementary materials.

## Acknowledgements

The authors have no additional acknowledgements to report.

## Ethics approval

Not applicable. This systematic review and meta-analysis was based exclusively on aggregate data obtained from previously published studies. No new human participants were recruited, no intervention was performed, no identifiable personal information was accessed, and no new individual-level participant data or biological material was collected. Therefore, approval from an institutional review board or research ethics committee was not required.

## Registration and reporting

The review was registered in PROSPERO under registration number CRD420261321165 and was conducted and reported in accordance with the PRISMA 2020 statement.

## Consent to participate

Not applicable. No human participants were recruited, contacted, or enrolled, and no new individual-level participant data were collected for this systematic review and meta-analysis.

## Consent for publication

Not applicable. This manuscript does not contain identifiable personal information, individual participant data, or identifiable clinical images.

## Data availability

All aggregate data supporting the findings of this systematic review and meta-analysis are included in the article and its supplementary materials. Supplementary Material 1 contains the complete database search strategies. Supplementary Material 2 contains the finalized decisions for all 154 full-text reports, including the 13 included studies and 141 exclusions grouped into the three PRISMA categories. Supplementary Material 3 contains the complete structured study-level data-extraction dataset in a single worksheet. Supplementary Material 4 contains the complete item-level Joanna Briggs Institute risk-of-bias assessments. Supplementary Material 5 contains the supplementary forest plots, subgroup and sensitivity analyses, influence diagnostics, and small-study-effect plots. Supplementary Material 6 contains the completed PRISMA 2020 checklist, indicating where each reporting item is addressed in the manuscript or supplementary materials.

## Author contributions

Farzan Fahim (FF) conceived the study, contributed to the development of the research question and methodology, co-designed the standardized data-extraction form, coordinated the study-selection and data-extraction procedures, participated in data verification, contributed to the interpretation of the findings, supervised preparation of the manuscript, and critically reviewed and revised the manuscript for important intellectual content.

Fatemeh Vosoughian (FV) conceived the study, contributed to development of the research question and methodology, participated in preparation of the protocol, designed the screening and exclusion forms, co-designed the standardized data-extraction form, coordinated the review procedures, contributed to interpretation of the findings, and participated in drafting and critical revision of the manuscript.

Amirmahdi Mojtahedzadeh (AMM) contributed to development of the methodology and analytical framework, adjudicated disagreements arising during study selection, data extraction, and methodological assessment, verified the extracted data, performed the statistical analyses, prepared the figures and tables, interpreted the findings, and drafted and critically revised the manuscript.

Barbod Mahdavi (BM) performed title and abstract screening according to the predefined eligibility criteria, contributed to verification of screening decisions, and critically reviewed and revised the manuscript.

Kiana Taghipoor (KT) performed title and abstract screening and full-text eligibility assessment, contributed to verification of study inclusion and exclusion decisions, and critically reviewed and revised the manuscript.

Rauf Rostami (RR) performed full-text eligibility assessment and data extraction, contributed to verification of the extracted study-level information, and critically reviewed and revised the manuscript.

Fatemeh Deldar (FD) performed title and abstract screening and data extraction, contributed to verification of the extracted data, and critically reviewed and revised the manuscript.

Ramtin Shemshadigolafzani (RSh) performed title and abstract screening according to the predefined eligibility criteria, contributed to verification of screening decisions, and critically reviewed and revised the manuscript.

Mahla Rakhshani (MR) and Reihane Qahremani (RQ) independently performed the Joanna Briggs Institute risk-of-bias assessments, contributed to resolution and verification of methodological judgments, and critically reviewed and revised the manuscript.

Fatemeh Gheibi (FG), Saeed-Rezaali (SRA), Nahal Badavi (NB), and Pardis fathabadi (PFA) contributed to verification and interpretation of the study-level findings and participated in the critical review, editing, and revision of the manuscript for important intellectual content.

Alireza Zali (AZ) provided senior supervision and neurosurgical expertise, contributed to interpretation of the findings and refinement of the clinical implications, and critically reviewed and revised the manuscript for important intellectual content.

FF and FV contributed equally to this work and share first authorship. FF is corresponding author. All authors made substantial contributions to the intellectual development of the work, reviewed and approved the final version of the manuscript, agreed to its submission, and accept accountability for all aspects of the work.

### Use of generative artificial intelligence

Generative AI was used only to refine language, readability, and formatting; all scientific content, analyses, interpretations, and conclusions were produced and verified by the authors.

## Supplementary material legends

Supplementary Material 1. Complete database search strategies for PubMed/MEDLINE, Embase, Scopus, Web of Science, and the Cochrane Library.

Supplementary Material 2. Final full-text screening workbook containing decisions for all 154 assessed reports: 13 included studies and 141 exclusions categorized as wrong study design (n = 16), no suitable outcomes (n = 56), or no eligible intervention or exposure (n = 69).

Supplementary Material 3. Single-worksheet study-level dataset containing characteristics, interventions or exposures, comparators, baseline status, outcomes, principal findings, and quantitative-synthesis inputs for the 13 included studies.

Supplementary Material 4. Complete item-level Joanna Briggs Institute risk-of-bias assessments for the 13 included studies.

Supplementary Material 5. Supplementary quantitative analyses, including exploratory overall models, subgroup and sensitivity analyses, leave-one-out and influence diagnostics, funnel plots, and trim-and-fill results.

Supplementary Material 6. Completed PRISMA 2020 checklist, indicating the location within the manuscript or supplementary materials where each reporting item is addressed.

