## Supplementary Material 1 for "Vitamin D Status and Supplementation in Patients Undergoing Spine Surgery: A Systematic Review and Meta-analysis"

### Supplementary Material 1. Complete database search strategies

Database-specific search strings, search date, and exported record counts used for the systematic review.

**Search date:** 1 June 2026

**Records exported:** PubMed/MEDLINE 141; Cochrane Library 132; Scopus 1,387; Embase 1,694; Web of Science 439 (total 3,793).

#### PubMed/MEDLINE

```
#1 AND #2 AND #3 AND #4
#1 AND #3 AND #4
(("spinal surgery"[Title/Abstract] OR ("Spinal Fusion"[MeSH Terms] OR "Spinal Fusions"[Title/Abstract] OR "Spondylodesis"[Title/Abstract] OR "Spondylodeses"[Title/Abstract] OR "Spondylosyndesis"[Title/Abstract]) OR ("lumbar vertebrae/surgery"[MeSH Terms] OR "Vertebrae surgery"[Title/Abstract]) OR "thoracic vertebrae/surgery"[MeSH Terms] OR ("Bone Screws"[MeSH Terms] OR "Bone Screw"[Title/Abstract]) OR "spine surgery"[Title/Abstract] OR "spinal fixation"[Title/Abstract] OR "lumbar fusion"[Title/Abstract] OR "thoracolumbar fusion"[Title/Abstract] OR ("posterior lumbar interbody fusion"[Title/Abstract] OR "PLIF"[Title/Abstract]) OR ("transforaminal lumbar interbody fusion"[Title/Abstract] OR "TLIF"[Title/Abstract]) OR ("anterior lumbar interbody fusion"[Title/Abstract] OR "ALIF"[Title/Abstract]) OR "Posterolateral Fusion"[Title/Abstract] OR "instrumented fusion"[Title/Abstract]) AND "randomized controlled trial"[Publication Type])
AND
(("Fusion Rate"[Title/Abstract] OR "Radiographic Fusion"[Title/Abstract] OR "Osseous Fusion"[Title/Abstract] OR "Bony Fusion"[Title/Abstract] OR "Solid Fusion"[Title/Abstract] OR "Incomplete Fusion"[Title/Abstract] OR "Partial Fusion"[Title/Abstract] OR "Nonunion"[Title/Abstract] OR ("Pseudarthrosis"[MeSH Terms] OR "Pseudarthrosis"[Title/Abstract]) OR "Fibrous Fusion"[Title/Abstract] OR "Bone Bridging"[Title/Abstract] OR "Trabecular Continuity"[Title/Abstract] OR "Intertransverse Fusion"[Title/Abstract] OR "Facet Fusion"[Title/Abstract] OR "Interfacetal Fusion"[Title/Abstract] OR "Interarticular Fusion"[Title/Abstract] OR "Posterolateral Fusion"[Title/Abstract] OR "Interbody Fusion"[Title/Abstract] OR "Anterior Fusion"[Title/Abstract] OR "Posterior Fusion"[Title/Abstract] OR "CT Fusion"[Title/Abstract] OR "Computed Tomography Fusion"[Title/Abstract] OR "Radiologic Assessment"[Title/Abstract] OR "Radiological Evaluation"[Title/Abstract] OR "Fusion Mass"[Title/Abstract] OR "Fusion Grading"[Title/Abstract] OR "Fusion Quality"[Title/Abstract] OR "Fusion Success"[Title/Abstract] OR "Fusion Failure"[Title/Abstract] OR "Lenke Criteria"[Title/Abstract] OR "Brantigan Steffee Classification"[Title/Abstract] OR "Bridwell Fusion Grading"[Title/Abstract] OR "Graft Incorporation"[Title/Abstract] OR ("Bone Remodeling"[MeSH Terms] OR "Bone Remodeling"[Title/Abstract]) OR "Bone Consolidation"[Title/Abstract] OR "Instrument Loosening"[Title/Abstract] OR "Screw Loosening"[Title/Abstract] OR "Implant Failure"[Title/Abstract] OR "Hardware Loosening"[Title/Abstract] OR "Graft Resorption"[Title/Abstract] OR ("Treatment Outcome"[MeSH Terms] OR "Clinical Outcome"[Title/Abstract] OR "Treatment Efficacy"[Title/Abstract] OR "Clinical Efficacy"[Title/Abstract] OR "Rehabilitation Outcome"[Title/Abstract] OR "Clinical Effectiveness"[Title/Abstract] OR "Treatment Effectiveness"[Title/Abstract] OR "Patient-Relevant Outcomes"[Title/Abstract] OR "Functional Recovery"[Title/Abstract] OR "Return to Function"[Title/Abstract] OR "Functional Outcome"[Title/Abstract] OR "Pain Improvement"[Title/Abstract]) OR "ODI"[Title/Abstract] OR ("Visual Analog Scale"[MeSH Terms] OR "Visual Analog Scales"[Title/Abstract] OR "VAS"[Title/Abstract]) OR "NRS"[Title/Abstract] OR ("Back Pain"[MeSH Terms] OR "Back Pains"[Title/Abstract] OR "Back Ache"[Title/Abstract] OR "Back Aches"[Title/Abstract] OR "Backache"[Title/Abstract] OR "Backaches"[Title/Abstract] OR "Back Pain with Radiation"[Title/Abstract] OR "Back Pain without Radiation"[Title/Abstract] OR "Vertebrogenic Pain Syndrome"[Title/Abstract] OR "Vertebrogenic Pain Syndromes"[Title/Abstract]) OR ("Low Back Pain"[MeSH Terms] OR "Low Back Pains"[Title/Abstract] OR "Low Back Ache"[Title/Abstract] OR "Low Back Aches"[Title/Abstract] OR "Low Backache"[Title/Abstract] OR "Lower Back Pain"[Title/Abstract] OR "Lower Back Pains"[Title/Abstract] OR "Lumbago"[Title/Abstract] OR "Mechanical Low Back Pain"[Title/Abstract] OR "Postural Low Back Pain"[Title/Abstract] OR "Recurrent Low Back Pain"[Title/Abstract]) OR "JOA"[Title/Abstract] OR ("Reoperation"[MeSH Terms] OR "Revision Surgery"[Title/Abstract] OR "Revision Surgeries"[Title/Abstract] OR "Repeat Surgery"[Title/Abstract] OR "Surgical Revision"[Title/Abstract] OR "Joint Revision"[Title/Abstract]) OR "Implant Stability"[Title/Abstract] OR "Hardware-Related Complication"[Title/Abstract] OR "Fusion Time"[Title/Abstract] OR ("Length of Stay"[MeSH Terms] OR "Stay Length"[Title/Abstract] OR "Hospital Stay"[Title/Abstract]) OR ("Bone Density"[MeSH Terms] OR "Bone Mineral Density"[Title/Abstract] OR "Bone Mineral Content"[Title/Abstract]) OR "BMD Change"[Title/Abstract] OR "Bone Strength"[Title/Abstract] OR "Bone Quality"[Title/Abstract] OR ("Physiologic Calcification"[Title/Abstract] OR "Bone Mineralization"[Title/Abstract]) OR "Bone Healing"[Title/Abstract] OR ("Osteogenesis"[MeSH Terms] OR "Ossification"[Title/Abstract] OR "Bone Formation"[Title/Abstract] OR "Osteoclastogenesis"[Title/Abstract]) OR ("Bone Regeneration"[MeSH Terms] OR "Osteoconduction"[Title/Abstract]) OR ("Bone Remodeling"[MeSH Terms] OR "Bone Turnover"[Title/Abstract])) AND "randomized controlled trial"[Publication Type])
AND
(("Vitamin D"[MeSH Terms] OR "Vitamin D"[Title/Abstract] OR "Calcium and Vitamin D Supplementation"[Title/Abstract] OR ("Cholecalciferol"[MeSH Terms] OR "Vitamin D3"[Title/Abstract] OR "Calciol"[Title/Abstract]) OR ("Calciferols"[Title/Abstract] OR "Vitamin D2"[Title/Abstract]) OR ("Calcitriol"[MeSH Terms] OR "1 25 dihydroxyvitamin d3"[Title/Abstract] OR "Rocaltrol"[Title/Abstract] OR "MC1288"[Title/Abstract] OR "Decostriol"[Title/Abstract]) OR "Alfacalcidol"[Title/Abstract] OR ("Hydroxycholecalciferols"[MeSH Terms] OR "Hydroxyvitamins D"[Title/Abstract]) OR ("Calcium"[MeSH Terms] OR "Calcium Supplementation"[Title/Abstract]) OR ("Calcium Carbonate"[MeSH Terms] OR "Calcite"[Title/Abstract]) OR "Calcium Citrate"[MeSH Terms] OR "Dietary Calcium"[Title/Abstract])))
```

#### Cochrane Library

```
( "spinal surgery":ti,ab OR "Spinal Fusion":ti,ab OR "Spinal Fusions":ti,ab OR "Spondylodesis":ti,ab OR "Spondylodeses":ti,ab OR "Spondylosyndesis":ti,ab OR "lumbar vertebrae surgery":ti,ab OR "Vertebrae surgery":ti,ab OR "thoracic vertebrae surgery":ti,ab OR "Bone Screws":ti,ab OR "Bone Screw":ti,ab OR "spine surgery":ti,ab OR "spinal fixation":ti,ab OR "lumbar fusion":ti,ab OR "thoracolumbar fusion":ti,ab OR "posterior lumbar interbody fusion":ti,ab OR "PLIF":ti,ab OR "transforaminal lumbar interbody fusion":ti,ab OR "TLIF":ti,ab OR "anterior lumbar interbody fusion":ti,ab OR "ALIF":ti,ab OR "Posterolateral Fusion":ti,ab OR "instrumented fusion":ti,ab OR [mh "Spinal Fusion"] OR [mh "Spinal Surgery"] ) AND ( "Fusion Rate":ti,ab OR "Radiographic Fusion":ti,ab OR "Osseous Fusion":ti,ab OR "Bony Fusion":ti,ab OR "Solid Fusion":ti,ab OR "Incomplete Fusion":ti,ab OR "Partial Fusion":ti,ab OR "Nonunion":ti,ab OR "Pseudarthrosis":ti,ab OR "Fibrous Fusion":ti,ab OR "Bone Bridging":ti,ab OR "Trabecular Continuity":ti,ab OR "Intertransverse Fusion":ti,ab OR "Facet Fusion":ti,ab OR "Interfacetal Fusion":ti,ab OR "Interarticular Fusion":ti,ab OR "Posterolateral Fusion":ti,ab OR "Interbody Fusion":ti,ab OR "Anterior Fusion":ti,ab OR "Posterior Fusion":ti,ab OR "CT Fusion":ti,ab OR "Computed Tomography
```

Fusion":ti,ab OR "Radiologic Assessment":ti,ab OR "Radiological Evaluation":ti,ab OR "Fusion Mass":ti,ab OR "Fusion Grading":ti,ab OR "Fusion Quality":ti,ab OR "Fusion Success":ti,ab OR "Fusion Failure":ti,ab OR "Lenke Criteria":ti,ab OR "Brantigan Steffee Classification":ti,ab OR "Bridwell Fusion Grading":ti,ab OR "Graft Incorporation":ti,ab OR "Bone Remodeling":ti,ab OR "Bone Consolidation":ti,ab OR "Instrument Loosening":ti,ab OR "Screw Loosening":ti,ab OR "Implant Failure":ti,ab OR "Hardware Loosening":ti,ab OR "Graft Resorption":ti,ab OR "Treatment Outcome":ti,ab OR "Clinical Outcome":ti,ab OR "Treatment Efficacy":ti,ab OR "Clinical Efficacy":ti,ab OR "Rehabilitation Outcome":ti,ab OR "Clinical Effectiveness":ti,ab OR "Treatment Effectiveness":ti,ab OR "Patient-Relevant Outcomes":ti,ab OR "Functional Recovery":ti,ab OR "Return to Function":ti,ab OR "Functional Outcome":ti,ab OR "Pain Improvement":ti,ab OR ODI:ti,ab OR "Visual Analog Scales":ti,ab OR VAS:ti,ab OR NRS:ti,ab OR "Back Pain":ti,ab OR "Back Pains":ti,ab OR "Back Ache":ti,ab OR "Back Aches":ti,ab OR Backache:ti,ab OR Backaches:ti,ab OR "Back Pain with Radiation":ti,ab OR "Back Pain without Radiation":ti,ab OR "Vertebrogenic Pain Syndrome":ti,ab OR "Vertebrogenic Pain Syndromes":ti,ab OR "Low Back Pain":ti,ab OR "Low Back Pains":ti,ab OR "Low Back Ache":ti,ab OR "Low Back Aches":ti,ab OR "Low Backache":ti,ab OR "Lower Back Pain":ti,ab OR "Lower Back Pains":ti,ab OR "Lumbago":ti,ab OR "Mechanical Low Back Pain":ti,ab OR "Postural Low Back Pain":ti,ab OR "Recurrent Low Back Pain":ti,ab OR JOA:ti,ab OR "Revision Surgery":ti,ab OR "Revision Surgeries":ti,ab OR "Repeat Surgery":ti,ab OR "Surgical Revision":ti,ab OR "Joint Revision":ti,ab OR "Implant Stability":ti,ab OR "Hardware-Related Complication":ti,ab OR "Fusion Time":ti,ab OR "Stay Length":ti,ab OR "Hospital Stay":ti,ab OR "Bone Mineral Density":ti,ab OR "Bone Mineral Content":ti,ab OR "BMD Change":ti,ab OR "Bone Strength":ti,ab OR "Bone Quality":ti,ab OR "Physiologic Calcification":ti,ab OR "Bone Mineralization":ti,ab OR "Bone Healing":ti,ab OR "Ossification":ti,ab OR "Bone Formation":ti,ab OR "Osteoclastogenesis":ti,ab OR "Osteoconduction":ti,ab OR "Bone Turnover":ti,ab ) AND ( "Vitamin D":ti,ab OR "Calcium and Vitamin D Supplementation":ti,ab OR "Vitamin D3":ti,ab OR "Calciol":ti,ab OR "Vitamin D2":ti,ab OR "1 25 dihydroxyvitamin d3":ti,ab OR "Rocaltrol":ti,ab OR "MC1288":ti,ab OR "Decostriol":ti,ab OR "Alfacalcidol":ti,ab OR "Hydroxyvitamins D":ti,ab OR "Calcium":ti,ab OR "Calcium Supplementation":ti,ab OR "Calcite":ti,ab OR "Calcium Citrate":ti,ab OR "Dietary Calcium":ti,ab OR [mh "Vitamin D"] OR [mh "Calcium"] OR [mh "Calcium and Vitamin D Supplementation"] )

#### Scopus

( TITLE-ABS-KEY ( "spinal surgery" OR "Spinal Fusion" OR "Spinal Fusions" OR "Spondylodesis" OR "Spondylodeses" OR "Spondylosyndesis" OR "lumbar vertebrae surgery" OR "Vertebrae surgery" OR "thoracic vertebrae surgery" OR "Bone Screws" OR "Bone Screw" OR "spine surgery" OR "spinal fixation" OR "lumbar fusion" OR "thoracolumbar fusion" OR "posterior lumbar interbody fusion" OR "PLIF" OR "transforaminal lumbar interbody fusion" OR "TLIF" OR "anterior lumbar interbody fusion" OR "ALIF" OR "Posterolateral Fusion" OR "instrumented fusion" ) ) AND ( TITLE-ABS-KEY ( "Fusion Rate" OR "Radiographic Fusion" OR "Osseous Fusion" OR "Bony Fusion" OR "Solid Fusion" OR "Incomplete Fusion" OR "Partial Fusion" OR "Nonunion" OR "Pseudarthrosis" OR "Fibrous Fusion" OR "Bone Bridging" OR "Trabecular Continuity" OR "Intertransverse Fusion" OR "Facet Fusion" OR "Interfacetal Fusion" OR "Interarticular Fusion" OR "Posterolateral Fusion" OR "Interbody Fusion" OR "Anterior Fusion" OR "Posterior Fusion" OR "CT Fusion" OR "Computed Tomography Fusion" OR "Radiologic Assessment" OR "Radiological Evaluation" OR "Fusion Mass" OR "Fusion Grading" OR "Fusion Quality" OR "Fusion Success" OR "Fusion Failure" OR "Lenke Criteria" OR "Brantigan Steffee Classification" OR "Bridwell Fusion Grading" OR "Graft Incorporation" OR "Bone Remodeling" OR "Bone Consolidation" OR "Instrument Loosening" OR "Screw Loosening" OR "Implant Failure" OR "Hardware Loosening" OR "Graft Resorption" OR "Treatment Outcome" OR "Clinical Outcome" OR "Treatment Efficacy" OR "Clinical Efficacy" OR "Rehabilitation Outcome" OR "Clinical Effectiveness" OR "Treatment Effectiveness" OR "Patient-Relevant Outcomes" OR "Functional Recovery" OR "Return to Function" OR "Functional Outcome" OR "Pain Improvement" OR "ODI" OR "Visual Analog Scale" OR "Visual Analog Scales" OR "VAS" OR "NRS" OR "Back Pain" OR "Back Pains" OR "Back Ache" OR "Back Aches" OR "Backache" OR "Backaches" OR "Back Pain with Radiation" OR "Back Pain without Radiation" OR "Vertebrogenic Pain Syndrome" OR "Vertebrogenic Pain Syndromes" OR "Low Back Pain" OR "Low Back Pains" OR "Low Back Ache" OR "Low Back Aches" OR "Low Backache" OR "Lower Back Pain" OR "Lower Back Pains" OR "Lumbago" OR "Mechanical Low Back Pain" OR "Postural Low Back Pain" OR "Recurrent Low Back Pain" OR "JOA" OR "Reoperation" OR "Revision Surgery" OR "Revision Surgeries" OR "Repeat Surgery" OR "Surgical Revision" OR "Joint Revision" OR "Implant Stability" OR "Hardware-Related Complication" OR "Fusion Time" OR "Length of Stay" OR "Stay Length" OR "Hospital Stay" OR "Bone Density" OR "Bone Mineral Density" OR "Bone Mineral Content" OR "BMD Change" OR "Bone Strength" OR "Bone Quality" OR "Physiologic Calcification" OR "Bone Mineralization" OR "Bone Healing" OR "Osteogenesis" OR "Ossification" OR "Bone Formation" OR "Osteoclastogenesis" OR "Bone Regeneration" OR "Osteoconduction" OR "Bone Turnover" ) ) AND ( TITLE-ABS-KEY ( "Vitamin D" OR "Calcium and Vitamin D Supplementation" OR "Cholecalciferol" OR "Vitamin D3" OR "Calciol" OR "Calciferols" OR "Vitamin D2" OR "Calcitriol" OR "1 25 dihydroxyvitamin d3" OR "Rocaltrol" OR "MC1288" OR "Decostriol" OR "Alfacalcidol" OR "Hydroxycholecalciferols" OR "Hydroxyvitamins D" OR "Calcium" OR "Calcium Supplementation" OR "Calcium Carbonate" OR "Calcite" OR "Calcium Citrate" OR "Dietary Calcium" ) )

#### Embase

#1 AND #3 :936

('spinal surgery':ti,ab OR 'spinal fusion'/exp OR 'spinal fusions':ti,ab OR 'spondylodesis':ti,ab OR 'spondylodeses':ti,ab OR 'spondylosyndesis':ti,ab OR 'lumbar vertebrae/surgery' OR 'vertebrae surgery':ti,ab OR 'thoracic vertebrae/surgery' OR 'bone screws'/exp OR 'bone screw':ti,ab OR 'spine surgery':ti,ab OR 'spinal fixation':ti,ab OR 'lumbar fusion':ti,ab OR 'thoracolumbar fusion':ti,ab OR 'posterior lumbar interbody fusion':ti,ab OR 'plif':ti,ab OR 'transforaminal lumbar interbody fusion':ti,ab OR 'tlif':ti,ab OR 'anterior lumbar interbody fusion':ti,ab OR 'alif':ti,ab OR 'posterolateral fusion':ti,ab OR 'instrumented fusion':ti,ab) AND ('fusion rate':ti,ab OR 'radiographic fusion':ti,ab OR 'osseous fusion':ti,ab OR 'pseudarthrosis'/exp OR 'fusion success':ti,ab OR 'fusion failure':ti,ab OR 'bone remodeling'/exp OR 'bone consolidation':ti,ab OR 'instrument loosening':ti,ab OR 'implant failure':ti,ab OR 'treatment outcome'/exp OR 'clinical outcome':ti,ab OR 'functional recovery':ti,ab OR 'back pain'/exp OR 'low back pain'/exp OR 'reoperation'/exp OR 'length of stay'/exp OR 'bone density'/exp OR 'bone mineral density':ti,ab OR 'bone healing':ti,ab OR 'osteogenesis'/exp OR 'bone regeneration'/exp)

AND

('vitamin d'/exp OR 'vitamin d':ti,ab OR 'calcium and vitamin d supplementation':ti,ab OR 'cholecalciferol'/exp OR 'vitamin d3':ti,ab OR 'calciferols':ti,ab OR 'calcitriol'/exp OR '1 25 dihydroxyvitamin d3':ti,ab OR 'rocaltrol':ti,ab OR 'alfacalcidol':ti,ab OR 'hydroxycholecalciferols'/exp OR 'calcium'/exp OR 'calcium supplementation':ti,ab OR 'calcium carbonate'/exp OR 'calcium citrate'/exp OR 'dietary calcium':ti,ab)

**Search history: #1 AND #2 AND #3 :634**

(('spinal surgery':ti,ab OR 'Spinal Fusion'/exp OR 'Spinal Fusions':ti,ab OR 'Spondylodesis':ti,ab OR 'Spondylodeses':ti,ab OR 'Spondylosyndesis':ti,ab OR 'lumbar vertebrae/surgery'/exp OR 'Vertebrae surgery':ti,ab OR 'thoracic vertebrae/surgery'/exp OR 'Bone Screws'/exp OR 'Bone Screw':ti,ab OR 'spine surgery':ti,ab OR 'spinal fixation':ti,ab OR 'lumbar fusion':ti,ab OR 'thoracolumbar fusion':ti,ab OR 'posterior lumbar interbody fusion':ti,ab OR 'PLIF':ti,ab OR 'transforaminal lumbar interbody fusion':ti,ab OR 'TLIF':ti,ab OR 'anterior lumbar interbody fusion':ti,ab OR 'ALIF':ti,ab OR 'Posterolateral Fusion':ti,ab OR 'instrumented fusion':ti,ab)

AND

('Fusion Rate':ti,ab OR 'Radiographic Fusion':ti,ab OR 'Osseous Fusion':ti,ab OR 'Pseudarthrosis'/exp OR 'Fusion Success':ti,ab OR 'Fusion Failure':ti,ab OR 'Bone Remodeling'/exp OR 'Bone Consolidation':ti,ab OR 'Instrument Loosening':ti,ab OR 'Implant Failure':ti,ab OR 'Treatment Outcome'/exp OR 'Clinical Outcome':ti,ab OR 'Functional Recovery':ti,ab OR 'Back Pain'/exp OR 'Low Back Pain'/exp OR 'Reoperation'/exp OR 'Length of Stay'/exp OR 'Bone Density'/exp OR 'Bone Mineral Density':ti,ab OR 'Bone Healing':ti,ab OR 'Osteogenesis'/exp OR 'Bone Regeneration'/exp)

AND

('Vitamin D'/exp OR 'Vitamin D':ti,ab OR 'Calcium and Vitamin D Supplementation':ti,ab OR 'Cholecalciferol'/exp OR 'Vitamin D3':ti,ab OR 'Calciferols':ti,ab OR 'Calcitriol'/exp OR '1 25 dihydroxyvitamin d3':ti,ab OR 'Rocaltrol':ti,ab OR 'Alfacalcidol':ti,ab OR 'Hydroxycholecalciferols'/exp OR 'Calcium'/exp OR 'Calcium Supplementation':ti,ab OR 'Calcium Carbonate'/exp OR 'Calcium Citrate'/exp OR 'Dietary Calcium':ti,ab))

#### Web of Science

TS=( ( "spinal surgery" OR "Spinal Fusion" OR "Spinal Fusions" OR "Spondylodesis" OR "Spondylodeses" OR "Spondylosyndesis" OR "lumbar vertebrae surgery" OR "Vertebrae surgery" OR "thoracic vertebrae surgery" OR "Bone Screws" OR "Bone Screw" OR "spine surgery" OR "spinal fixation" OR "lumbar fusion" OR "thoracolumbar fusion" OR "posterior lumbar interbody fusion" OR "PLIF" OR "transforaminal lumbar interbody fusion" OR "TLIF" OR "anterior lumbar interbody fusion" OR "ALIF" OR "Posterolateral Fusion" OR "instrumented fusion" ) AND ( "Fusion Rate" OR "Radiographic Fusion" OR "Osseous Fusion" OR "Bony Fusion" OR "Solid Fusion" OR "Incomplete Fusion" OR "Partial Fusion" OR "Nonunion" OR "Pseudarthrosis" OR "Fibrous Fusion" OR "Bone Bridging" OR "Trabecular Continuity" OR "Intertransverse Fusion" OR "Facet Fusion" OR "Interfacetal Fusion" OR "Interarticular Fusion" OR "Interbody Fusion" OR "Anterior Fusion" OR "Posterior Fusion" OR "CT Fusion" OR "Computed Tomography Fusion" OR "Radiologic Assessment" OR "Radiological Evaluation" OR "Fusion Mass" OR "Fusion Grading" OR "Fusion Quality" OR "Fusion Success" OR "Fusion Failure" OR "Lenke Criteria" OR "Brantigan Steffee Classification" OR "Bridwell Fusion Grading" OR "Graft Incorporation" OR "Bone Remodeling" OR "Bone Consolidation" OR "Instrument Loosening" OR "Screw Loosening" OR "Implant Failure" OR "Hardware Loosening" OR "Graft Resorption" OR "Treatment Outcome" OR "Clinical Outcome" OR "Treatment Efficacy" OR "Clinical Efficacy" OR "Rehabilitation Outcome" OR "Clinical Effectiveness" OR "Treatment Effectiveness" OR "Patient-Relevant Outcomes" OR "Functional Recovery" OR "Return to Function" OR "Functional Outcome" OR "Pain Improvement" OR ODI OR "Visual Analog Scale" OR "Visual Analog Scales" OR VAS OR NRS OR "Back Pain" OR "Back Pains" OR "Back Ache" OR "Back Aches" OR Backache OR Backaches OR "Low Back Pain" OR "Low Back Pains" OR "Low Back Ache" OR "Low Back Aches" OR "Lower Back Pain" OR "Lower Back Pains" OR "Lumbago" OR "Mechanical Low Back Pain" OR "Postural Low Back Pain" OR "Recurrent Low Back Pain" OR JOA OR "Reoperation" OR "Revision Surgery" OR "Revision Surgeries" OR "Repeat Surgery" OR "Surgical Revision" OR "Joint Revision" OR "Implant Stability" OR "Hardware-Related Complication" OR "Fusion Time" OR "Length of Stay" OR "Hospital Stay" OR "Hospital Stays" OR "Bone Density" OR "Bone Mineral Density" OR "Bone Mineral Content" OR "BMD Change" OR "Bone Strength" OR "Bone Quality" OR "Physiologic Calcification" OR "Bone Mineralization" OR "Bone Healing" OR Osteogenesis OR Ossification OR "Bone Formation" OR Osteoclastogenesis OR "Bone Regeneration" OR Osteoconduction OR "Bone Turnover" ) AND ( "Vitamin D" OR "Calcium and Vitamin D Supplementation" OR Cholecalciferol OR "Vitamin D3" OR Calciol OR Calciferols OR "Vitamin D2" OR Calcitriol OR "1 25 dihydroxyvitamin d3" OR Rocaltrol OR MC1288 OR Decostriol OR Alfacalcidol OR Hydroxycholecalciferols OR "Hydroxyvitamins D" OR Calcium OR "Calcium Supplementation" OR "Calcium Carbonate" OR Calcite OR "Calcium Citrate" OR "Dietary Calcium" ) )
