## Supplementary Material 4 for "Vitamin D Status and Supplementation in Patients Undergoing Spine Surgery: A Systematic Review and Meta-analysis"

| Randomized controlled trials | Q1 | Q2 | Q3 | Q4 | Q5 | Q6 | Q7 | Q8 | Q9 | Q10 | Q11 | Q12 | Q13 | Overall risk |
|---|---|---|---|---|---|---|---|---|---|---|---|---|---|---|
| Placebo-controlled Trial of Oral Vitamin D and Alendronate Efficacy for Pain and Modic Changes in Lumbar Fusion Surgery | Yes | Yes | Yes | Yes | Unclear | Yes | Yes | Yes | Yes | Yes | Unclear | Yes | Unclear | Moderate |
| The efficacy of oral vitamin D supplements on fusion outcome in patients receiving elective lumbar spinal fusion—a randomized control trial | Yes | Unclear | Yes | Yes | Yes | Unclear | Yes | Yes | Yes | Yes | Yes | Yes | Unclear | Low |
| The Preoperative Supplementation With Vitamin D Attenuated Pain Intensity and Reduced the Level of Pro-inflammatory Markers in Patients After Posterior Lumbar Interbody Fusion | Unclear | Unclear | Yes | Yes | Yes | Unclear | Yes | Yes | Unclear | Yes | Yes | Yes | Unclear | Moderate |
| Comparing the Effect of Alendronate and Vitamin D Administration on Lumbosacral Fusion and Severity of Low Back Pain in Patients After Posterior Lumbar Fusion Surgery: A Randomized Clinical Trial | Yes | Unclear | No | Yes | Unclear | Yes | Yes | Yes | No | Yes | Yes | Yes | Unclear | Moderate |
| Early Rehabilitation Program and Vitamin D Supplementation Improves Sensitivity of Balance and the Postural Control in Patients after Posterior Lumbar Interbody Fusion: A Randomized Trial | Unclear | Yes | Unclear | Yes | Yes | Unclear | Yes | No | Unclear | Yes | Yes | Yes | Unclear | High |
| Vitamin D Supplementation Improves the Efects of the Rehabilitation Program on Balance and Pressure Distribution in Patients after Anterior Cervical Interbody Fusion-Randomized Control Trial | Yes | Unclear | No | Yes | Yes | Yes | Yes | No | Unclear | Yes | Yes | Yes | Unclear | Moderate |
| Vitamin D Deficiency Is Associated with Muscle Atrophy and Reduced Mitochondrial Function in Patients with Chronic Low Back Pain | Unclear | Unclear | Unclear | Yes | Unclear | Yes | Yes | Unclear | No | Yes | Yes | Yes | Unclear | High |

*Supplementary Table S1. JBI item-level assessments for randomized controlled trials; green indicates Yes, yellow Unclear, and red No.*

| Cohort studies | Q1 | Q2 | Q3 | Q4 | Q5 | Q6 | Q7 | Q8 | Q9 | Q10 | Q11 | |
|---|---|---|---|---|---|---|---|---|---|---|---|---|
| The effectiveness of vitamin D supplementation in functional outcome and quality of life (QoL) of lumbar spinal stenosis (LSS) requiring surgery | Yes | Yes | Yes | Yes | No | Yes | Unclear | Yes | Unclear | No | Yes | Moderate |
| Evaluation of the efficacy of combined vitamin D3 and K2 therapy in reducing implant-associated complication risk and improving spinal fusion stability | Yes | Yes | Yes | Yes | Yes | Yes | Yes | Yes | Yes | No | Unclear | Low |
| Effects of anti-osteoporosis treatment in elderly patients with osteoporosis and lumbar discectomy and fusion | No | Yes | Yes | No | No | Yes | Yes | Yes | Unclear | Unclear | No | High |
| Effects of 1, 25-Dihydroxyvitamin D3 on Posterior Transforaminal Lumbar Interbody Fusion in Patients with Osteoporosis and Lumbar Disc Degenerative Disease | Yes | Yes | Unclear | No | No | Yes | Yes | Yes | Unclear | Unclear | Unclear | Moderate |
| Association between antiosteoporosis medications and risk of sacral fracture after lumbosacral fusion in adults with osteoporosis: A proportional hazards analysis | Yes | Yes | Yes | Yes | No | Yes | Unclear | Yes | Unclear | No | Unclear | Moderate |

*Supplementary Table S2. JBI item-level assessments for cohort and case-control studies; green indicates Yes, yellow Unclear, and red No.*
