## Supplementary Material 5 for "Vitamin D Status and Supplementation in Patients Undergoing Spine Surgery: A Systematic Review and Meta-analysis"

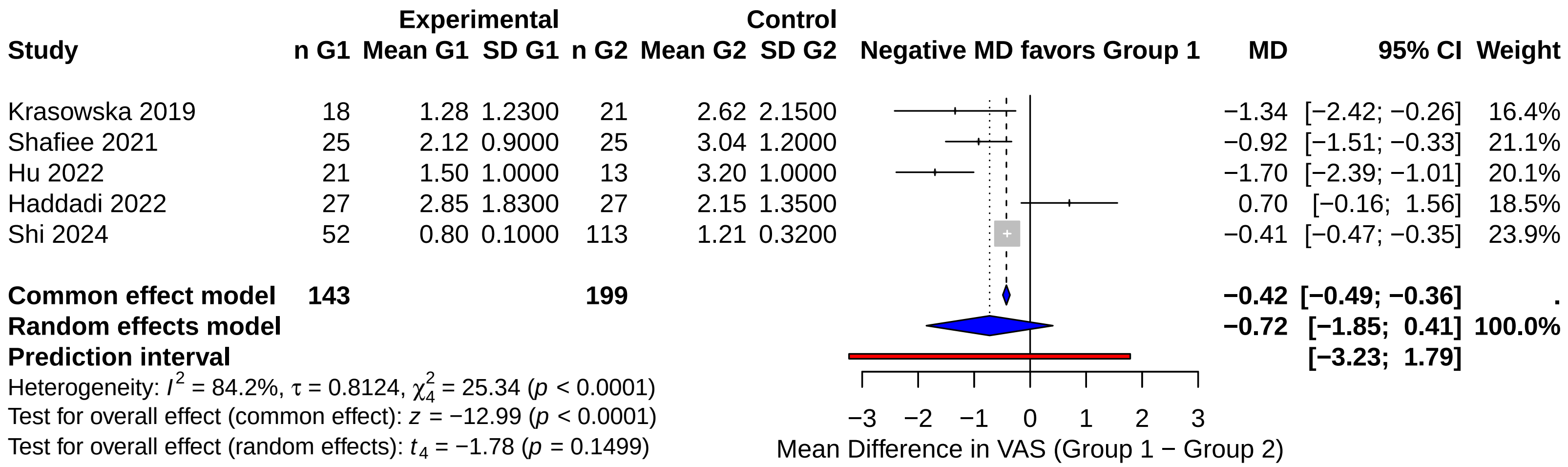


Fig. S1. Exploratory overall forest plot for postoperative VAS; study-specific and pooled mean differences are shown.

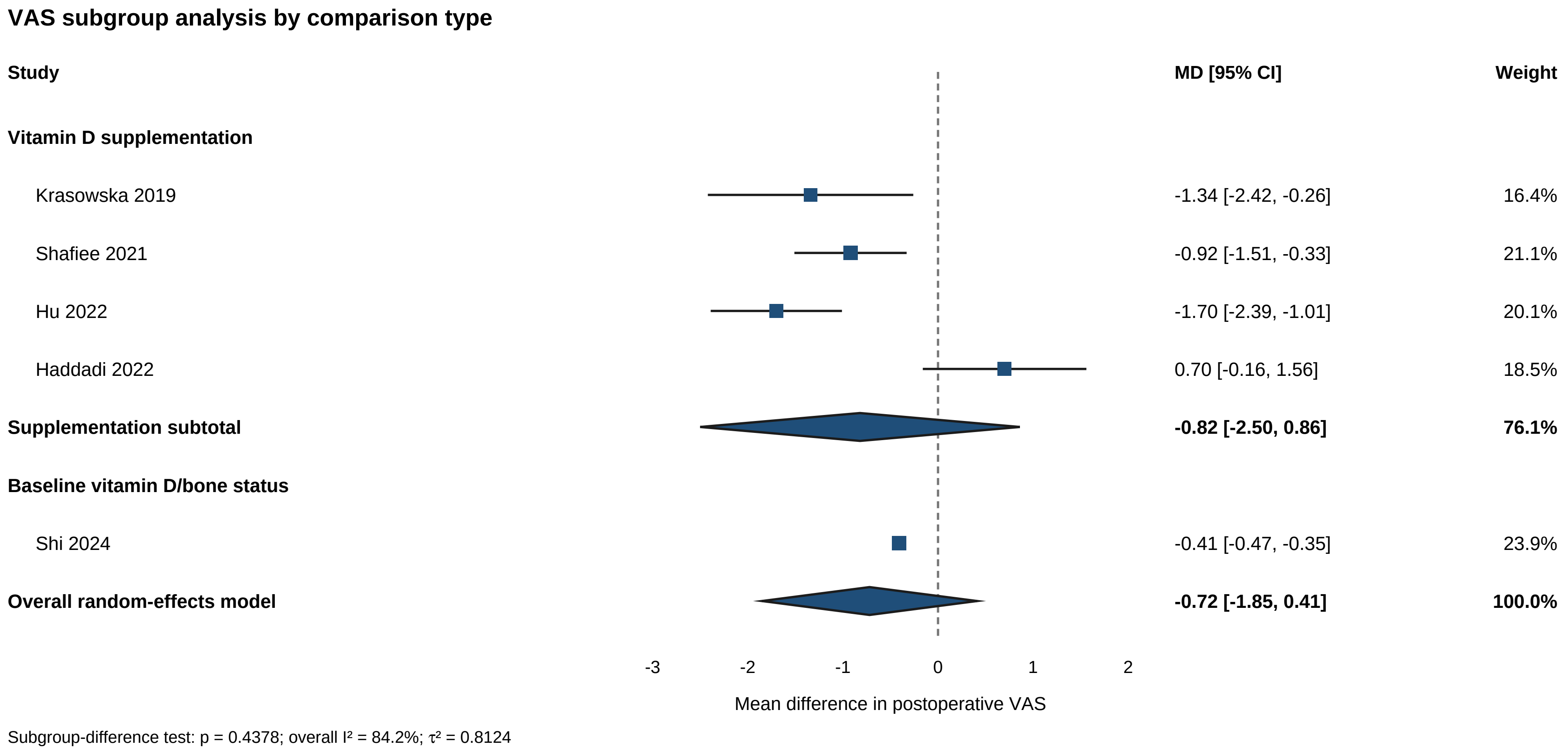


Fig. S2. VAS subgroup analysis by comparison type; direct supplementation and baseline-status contrasts are separated.

| Study | Experimental n G1 | Mean G1 | SD G1 | Control n G2 | Mean G2 | SD G2 |
|---|---|---|---|---|---|---|
| **Intervention_timing = Preoperative** | | | | | | |
| Krasowska 2019 | 18 | 1.28 | 1.23 | 21 | 2.62 | 2.15 |
| **Intervention_timing = Postoperative** | | | | | | |
| Hu 2022 | 21 | 1.50 | 1.00 | 13 | 3.20 | 1.00 |
| Haddadi 2022 | 27 | 2.85 | 1.83 | 27 | 2.15 | 1.35 |
| **Total (common effect, 95% CI)** | **48** | | | **40** | | |
| **Total (random effect, 95% CI)** | | | | | | |
| Heterogeneity: $Tau^2 = 2.7220$; $Chi^2 = 18.22$, df = 1 ($P < 0.0001$); $I^2 = 94.5\%$ | | | | | | |
| **Intervention_timing = No supplementation; baseline status** | | | | | | |
| Shi 2024 | 52 | 0.80 | 0.10 | 113 | 1.21 | 0.32 |
| **Intervention_timing = Postoperative/unclear** | | | | | | |
| Shafiee 2021 | 25 | 2.12 | 0.90 | 25 | 3.04 | 1.20 |
| **Total (common effect, 95% CI)** | **143** | | | **199** | | |
| **Total (random effect, 95% CI)** | | | | | | |

Heterogeneity: $Tau^2 = 0.6601$; $Chi^2 = 25.34$, df = 4 ($P < 0.0001$); $I^2 = 84.2\%$
Test for overall effect (random effects): $t_4 = -1.78$ ($P = 0.1499$)
Test for subgroup differences (common effect): $Chi^2 = 7.12$, df = 3 ($P = 0.0683$)
Test for subgroup differences (random effects): $Chi^2 = 5.66$, df = 3 ($P = 0.1295$)

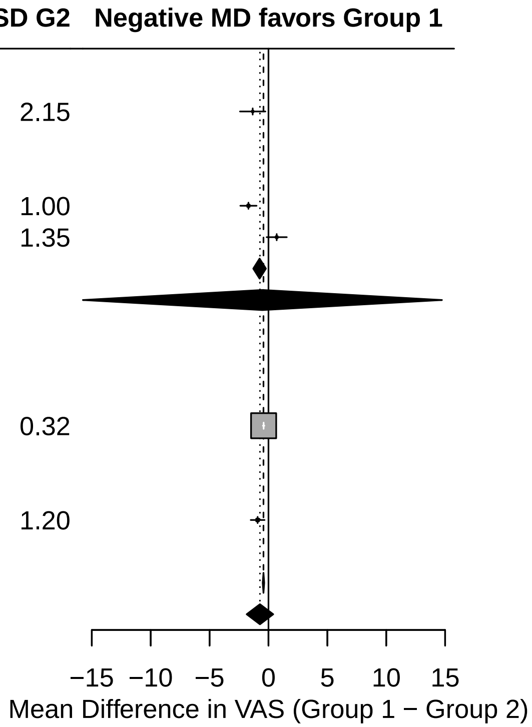


Fig. S3. VAS subgroup analysis by intervention timing; pooled effects are displayed for the prespecified timing groups.

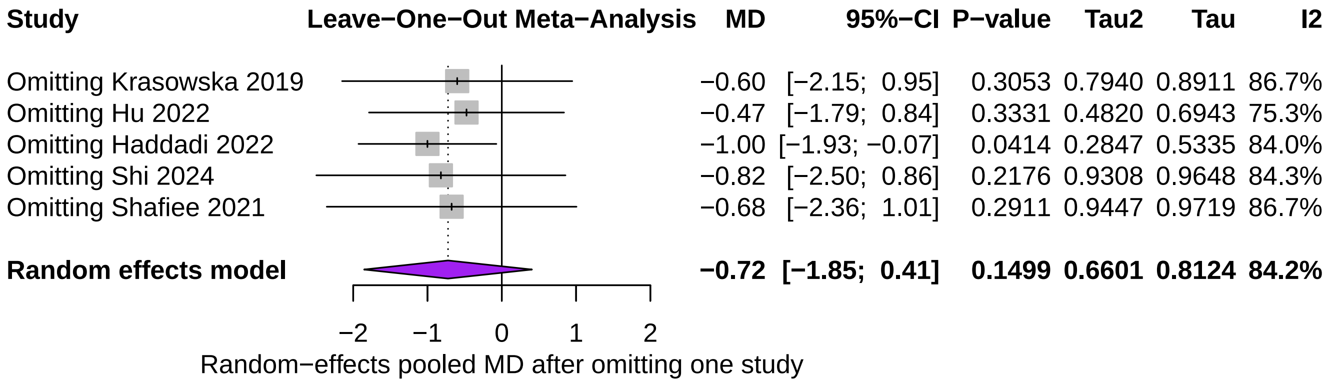


Fig. S4. Leave-one-out analysis for VAS; the pooled estimate is recalculated after omitting each study in turn.

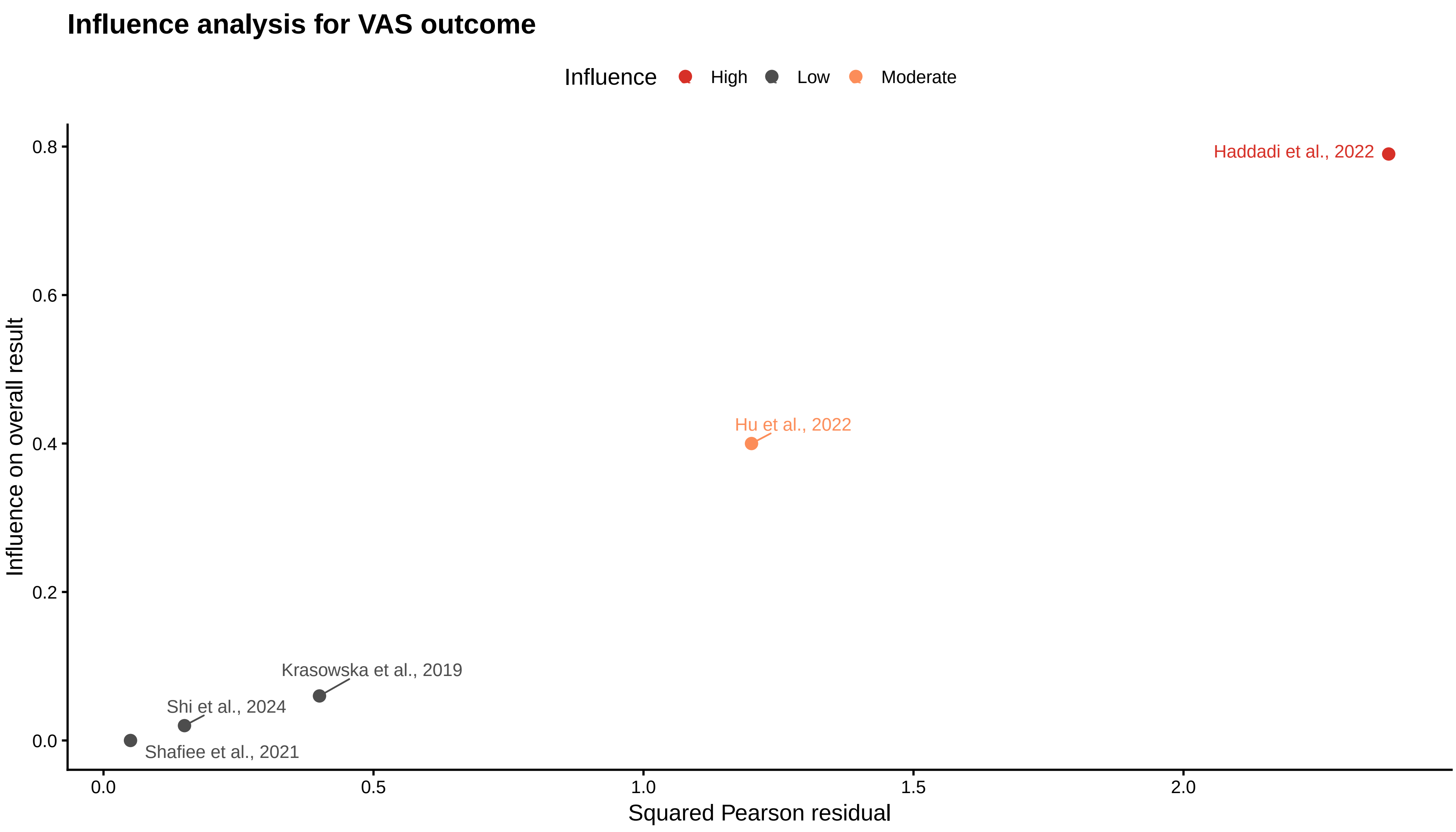


Fig. S5. Influence diagnostics for VAS; the plot identifies studies with the greatest influence on the pooled model.

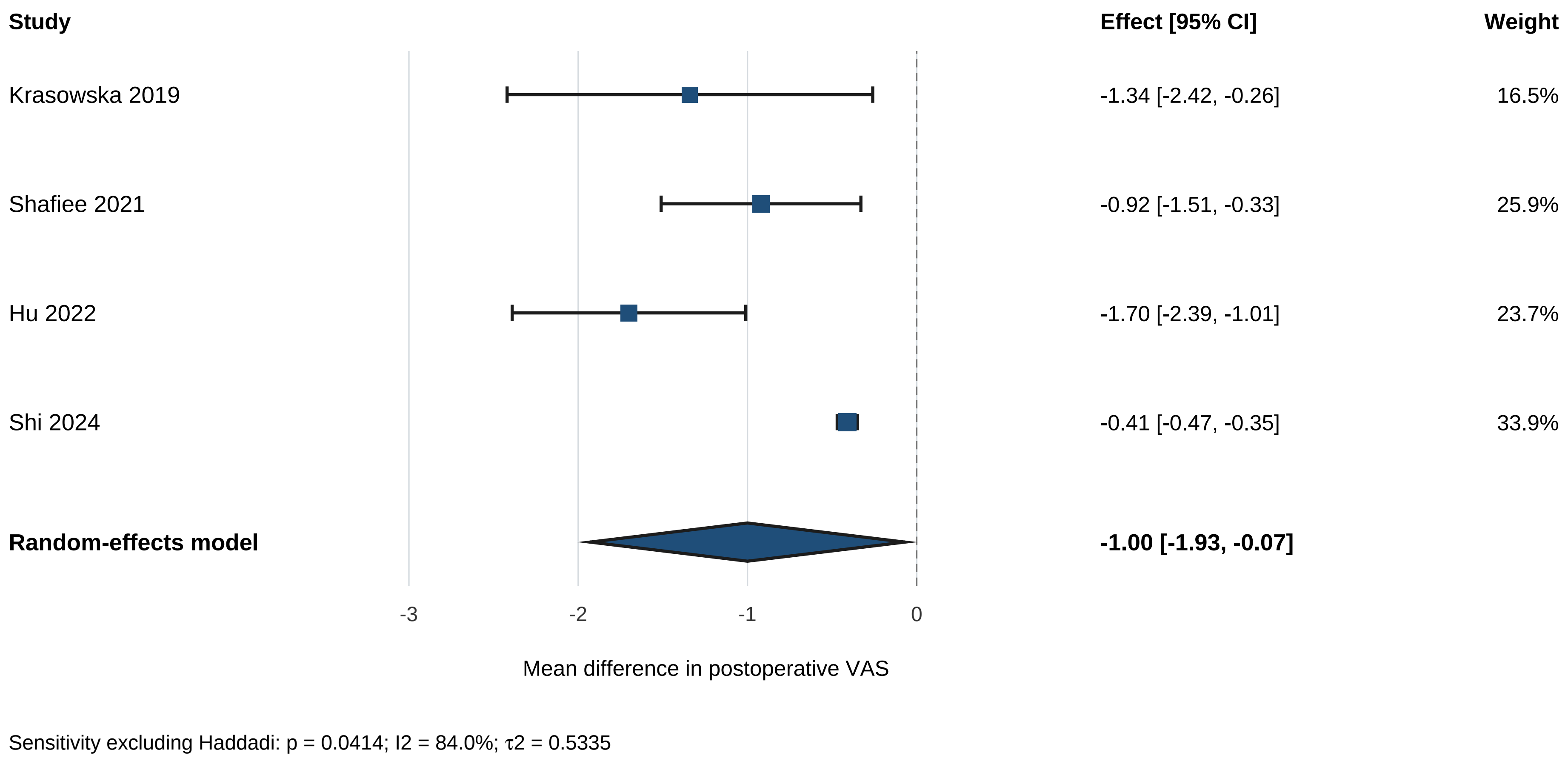


Fig. S6. VAS sensitivity analysis excluding Haddadi et al.; the pooled estimate is shown after removal of this study.

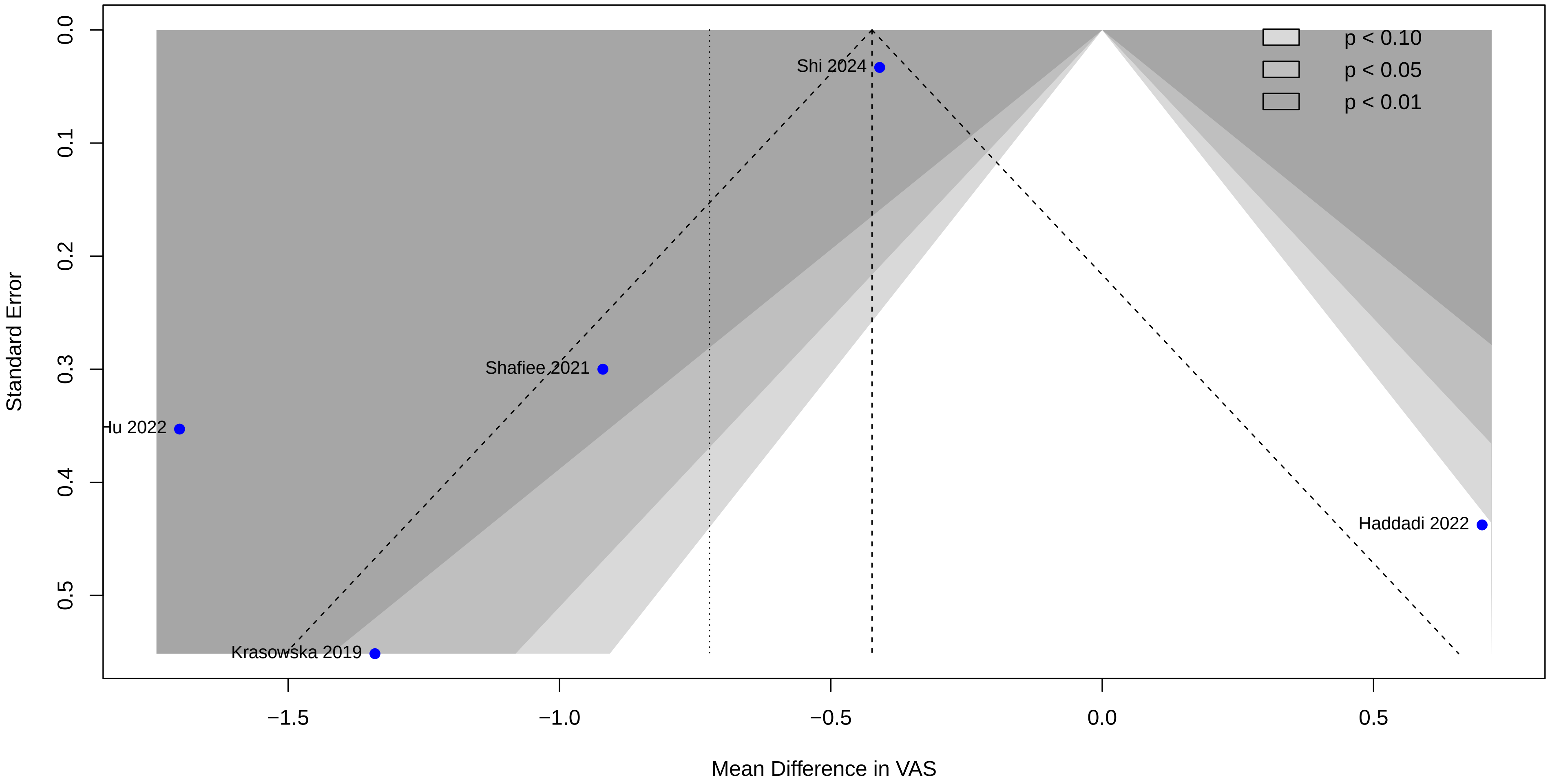


Fig. S7. Funnel plot for VAS; visual small-study-effect assessment is exploratory because few studies contributed.

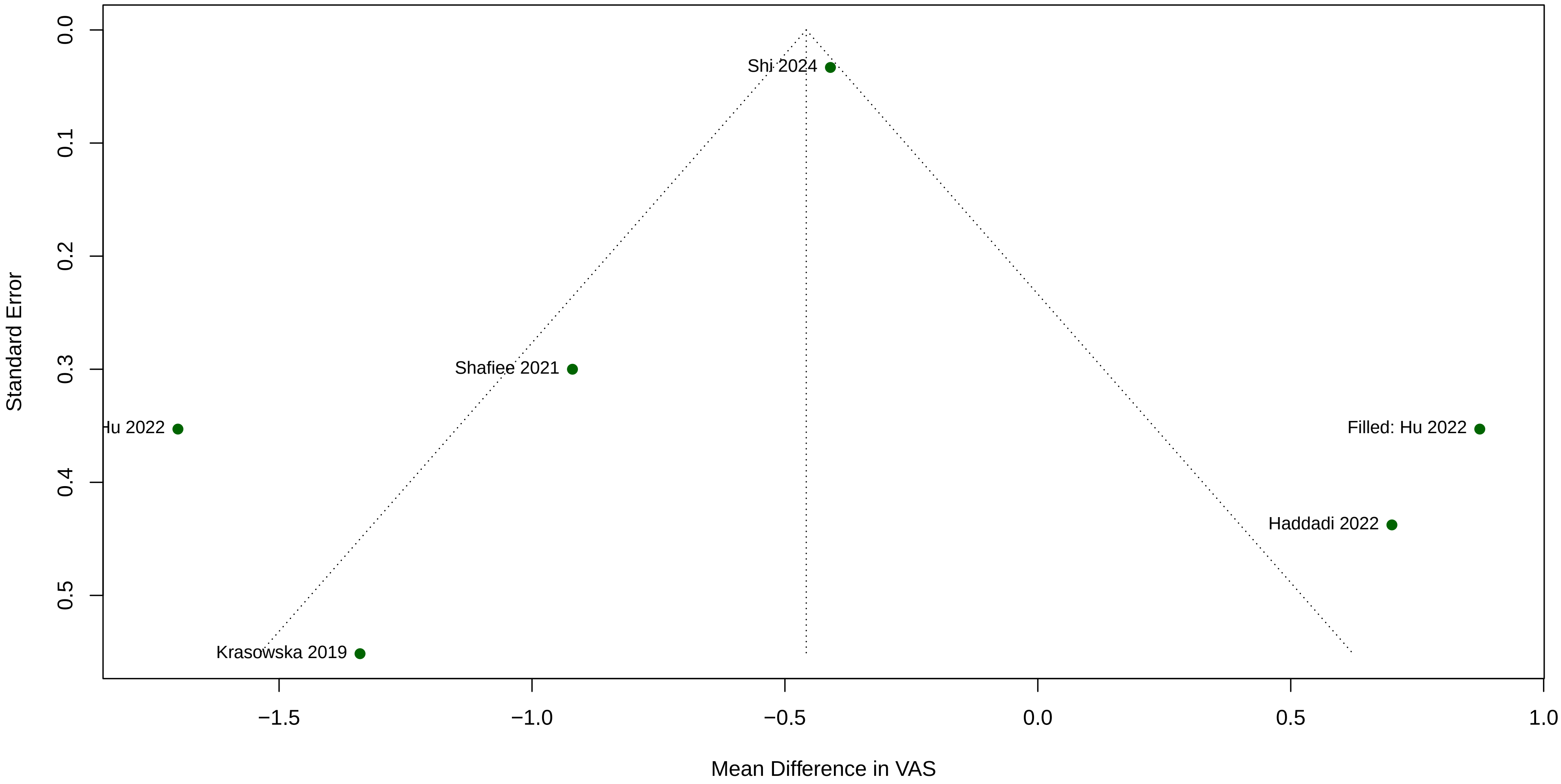


Fig. S8. Exploratory trim-and-fill analysis for VAS; the adjusted pattern is descriptive because the dataset is small.

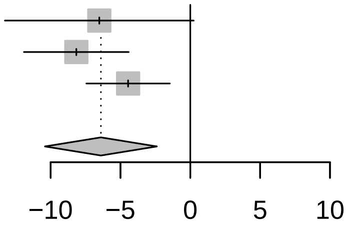


| Study | MD | 95% CI | P−value | Tau2 | Tau | I2 |
|---|---|---|---|---|---|---|
| Omitting Hu 2022 | −6.52 | [−13.28; 0.25] | 0.0589 | 23.4099 | 4.8384 | 98.3% |
| Omitting Shi 2024 | −8.16 | [−11.91; −4.41] | < 0.0001 | 6.6200 | 2.5729 | 89.8% |
| Omitting Ko 2020 | −4.45 | [ −7.44; −1.46] | 0.0035 | 3.7476 | 1.9359 | 80.0% |
| **Random effects model** | **−6.40** | **[−10.41; −2.39]** | **0.0017** | **11.8648** | **3.4445** | **96.8%** |

Fig. S9. Leave-one-out analysis for ODI; the pooled disability estimate is recalculated after each study is omitted.

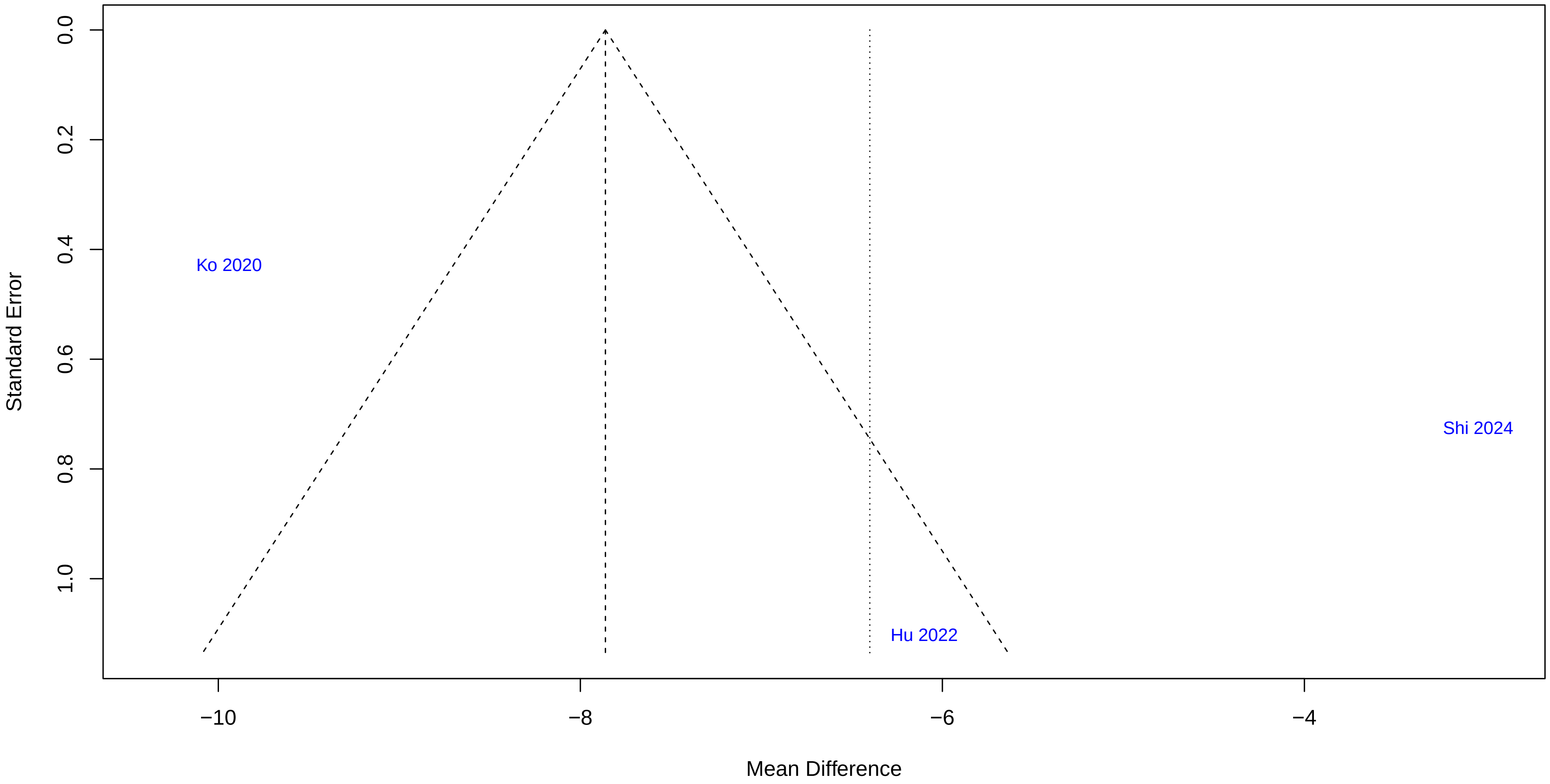


Fig. S10. Funnel plot for ODI; visual small-study-effect assessment is exploratory because only three studies contributed.

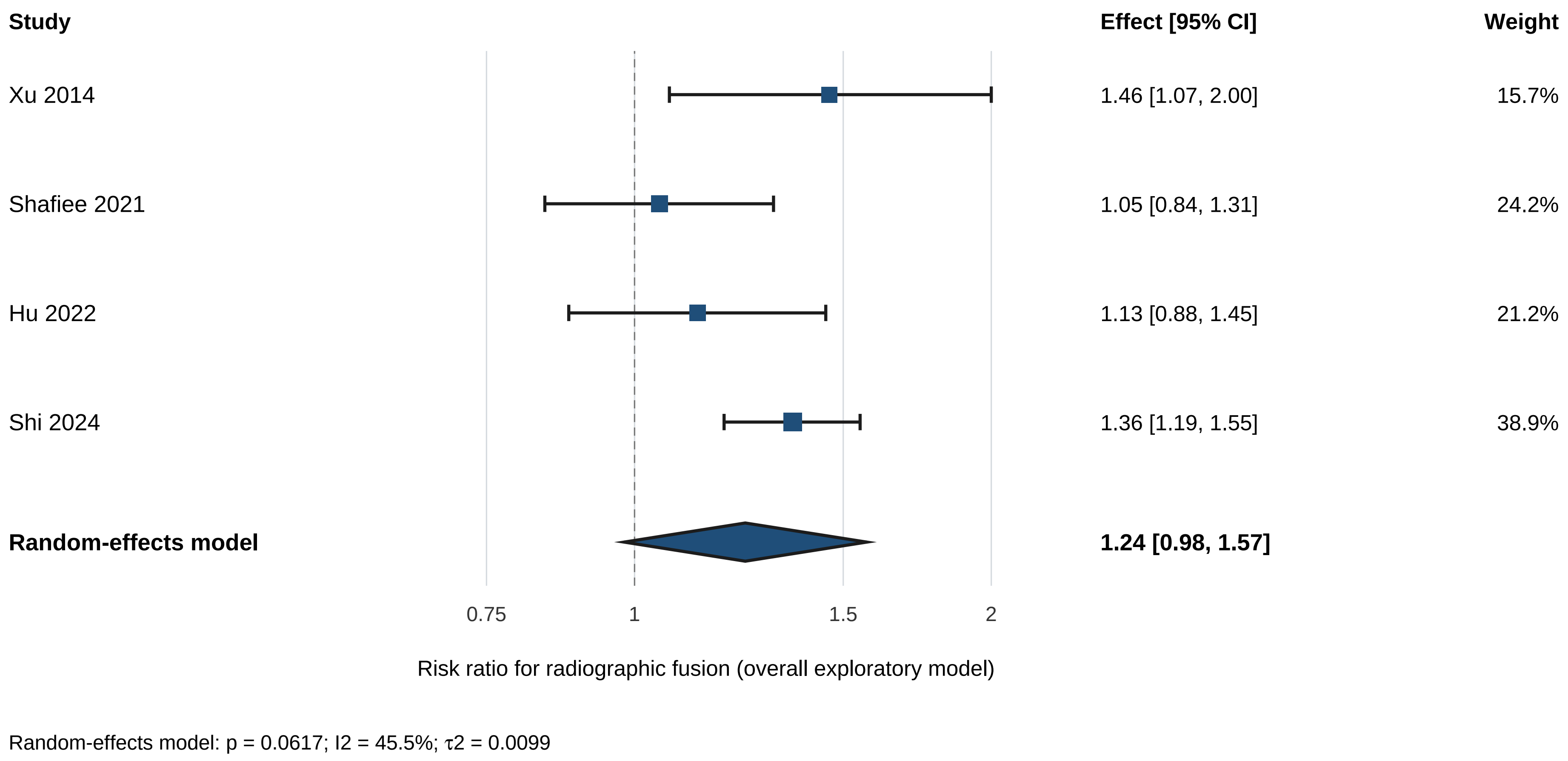


Fig. S11. Exploratory overall forest plot for fusion rate; study-specific and pooled risk ratios are shown.

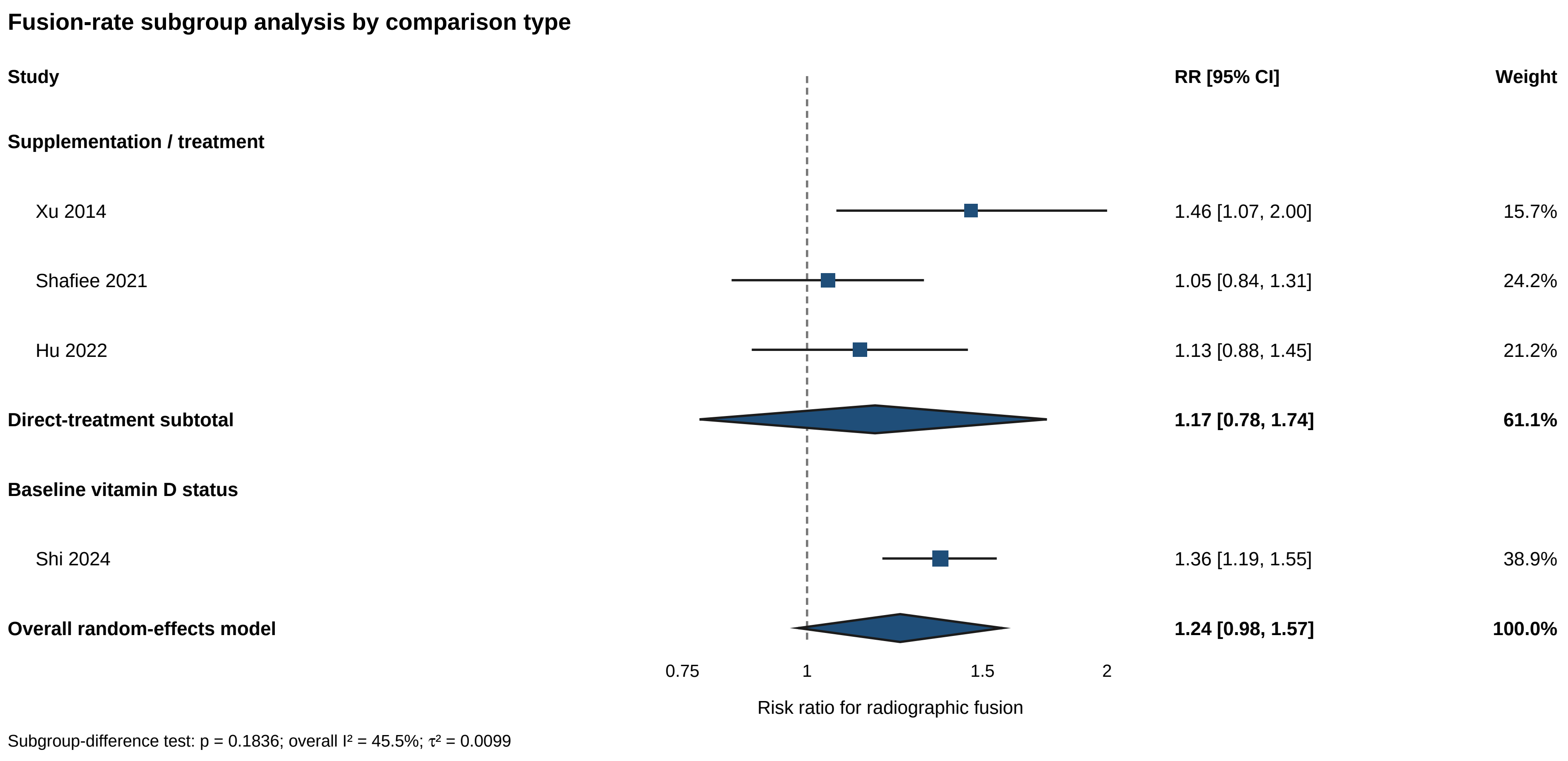


Fig. S12. Fusion-rate subgroup analysis by comparison type; direct treatment and baseline-status contrasts are separated.

### Leave-one-out analysis: overall fusion-rate dataset

| Omitted study | RR [95% CI] | p value | $\tau^2$ | $I^2$ |
|---|---|---|---|---|
| Hu 2022 | 1.27 [0.84, 1.91] | 0.1286 | 0.0148 | 55.7% |
| Shi 2024 | 1.17 [0.78, 1.74] | 0.2436 | 0.0062 | 31.5% |
| Shafiee 2021 | 1.32 [1.03, 1.69] | 0.0394 | <0.0001 | 6.3% |
| Xu 2014 | 1.20 [0.84, 1.70] | 0.1575 | 0.0127 | 56.6% |
| **Random-effects model** | **1.24 [0.98, 1.57]** | **0.0617** | **0.0099** | **45.5%** |

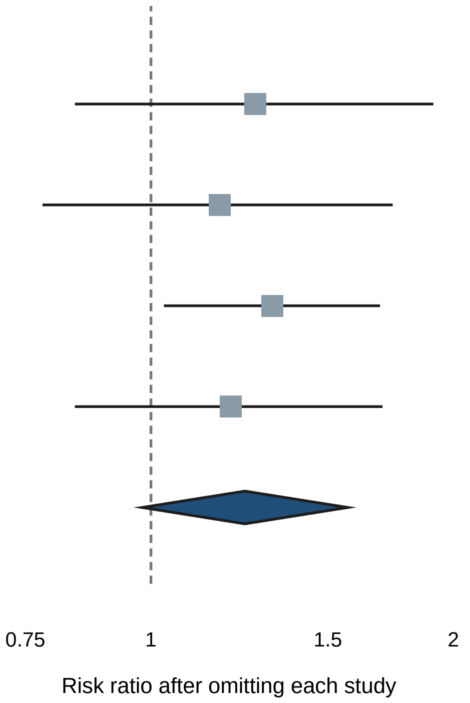


Fig. S13. Leave-one-out analysis for the overall fusion-rate dataset; each study is omitted sequentially.

### Leave-one-out analysis: supplementation-only fusion studies

| Omitted study | RR [95% CI] | p value | $\tau^2$ | $I^2$ |
|---|---|---|---|---|
| Hu 2022 | 1.21 [0.15, 9.88] | 0.4490 | 0.0358 | 65.0% |
| Shafiee 2021 | 1.26 [0.24, 6.49] | 0.3245 | 0.0129 | 38.1% |
| Xu 2014 | 1.08 [0.69, 1.70] | 0.2717 | 0 | 0% |
| **Random-effects model** | **1.17 [0.78, 1.74]** | **0.2436** | **0.0062** | **31.5%** |

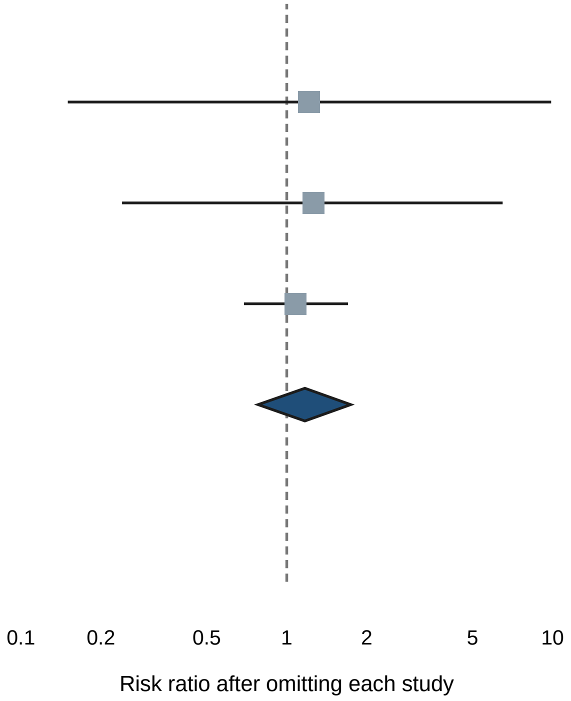


Fig. S14. Leave-one-out analysis for supplementation-only fusion studies; robustness to each study is displayed.

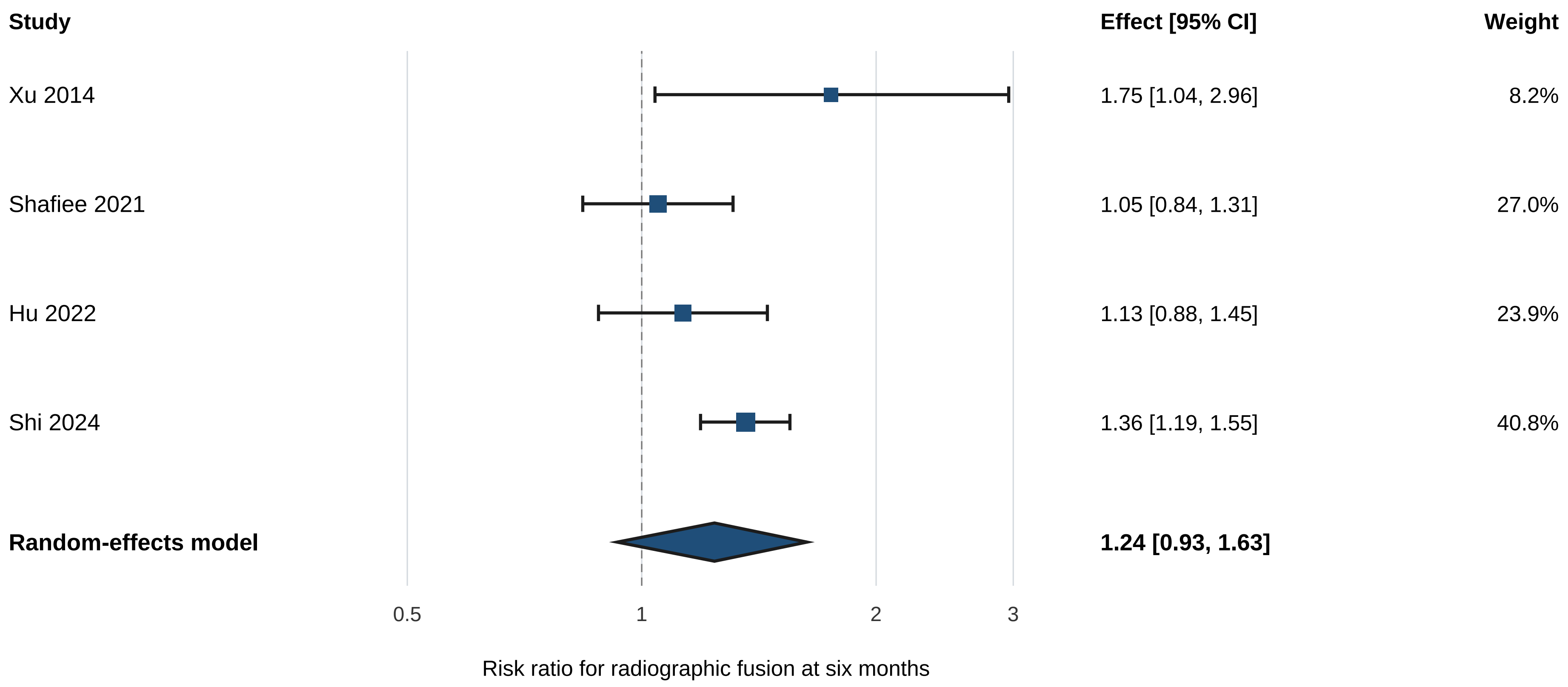


Sensitivity model: I2 = 51.5%; τ2 = 0.0125; heterogeneity p = 0.1031

Fig. S15. Six-month fusion-rate sensitivity analysis; pooled risk ratios are restricted to the six-month assessment.

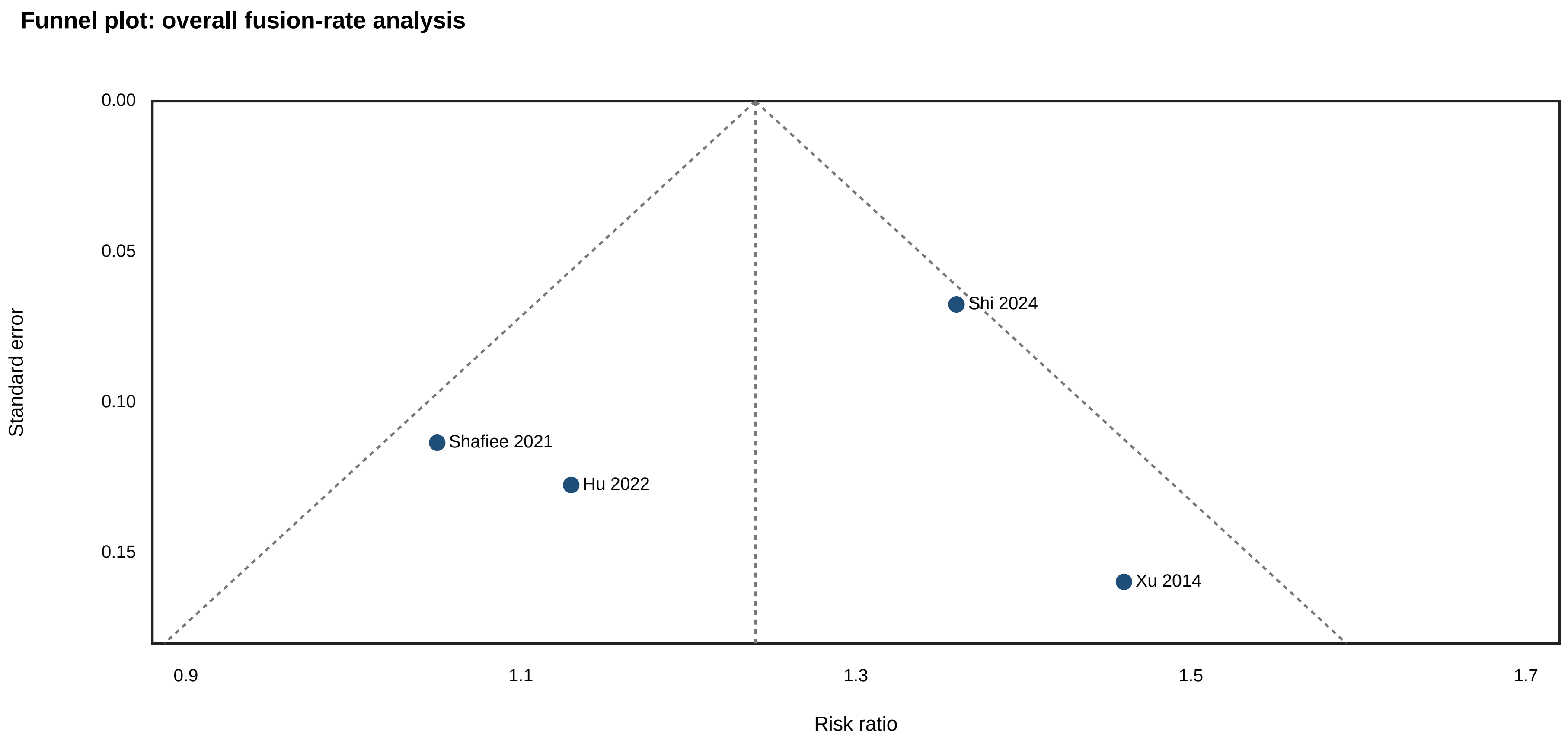


Fig. S16. Funnel plot for the overall fusion-rate analysis; interpretation is exploratory because few studies contributed.

### Funnel plot: supplementation-only fusion studies

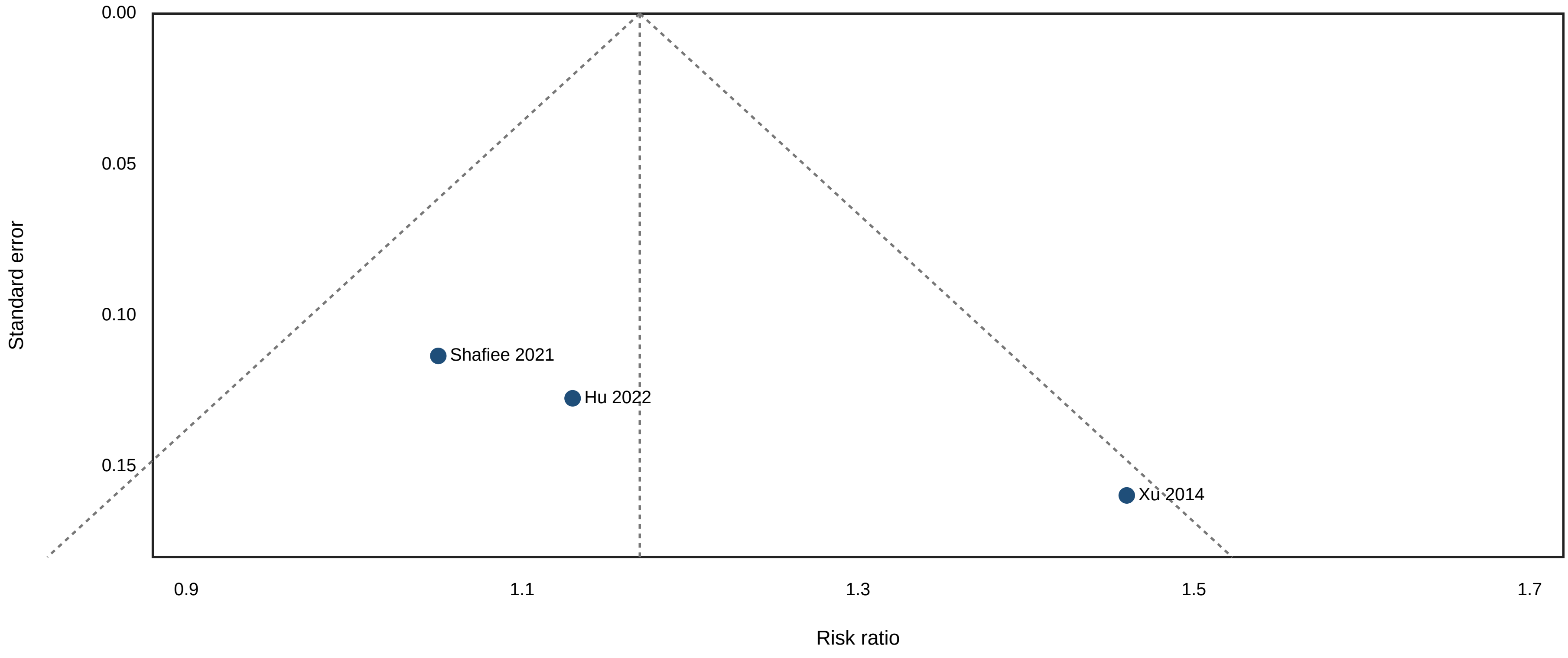


Fig. S17. Funnel plot for supplementation-only fusion studies; interpretation is exploratory because only three studies contributed.
