## Supplementary Material 6 for "Vitamin D Status and Supplementation in Patients Undergoing Spine Surgery: A Systematic Review and Meta-analysis"

| **Section and Topic** | **Item #** | **Checklist item** | **Location where item is reported** |
| --- | --- | --- | --- |
| **TITLE** | | |  |
| Title | 1 | Identify the report as a systematic review. |  |
| **ABSTRACT** | | |  |
| Abstract | 2 | See the PRISMA 2020 for Abstracts checklist. |  |
| **INTRODUCTION** | | |  |
| Rationale | 3 | Describe the rationale for the review in the context of existing knowledge. |  |
| Objectives | 4 | Provide an explicit statement of the objective(s) or question(s) the review addresses. |  |
| **METHODS** | | |  |
| Eligibility criteria | 5 | Specify the inclusion and exclusion criteria for the review and how studies were grouped for the syntheses. |  |
| Information sources | 6 | Specify all databases, registers, websites, organisations, reference lists and other sources searched or consulted to identify studies. Specify the date when each source was last searched or consulted. |  |
| Search strategy | 7 | Present the full search strategies for all databases, registers and websites, including any filters and limits used. |  |
| Selection process | 8 | Specify the methods used to decide whether a study met the inclusion criteria of the review, including how many reviewers screened each record and each report retrieved, whether they worked independently, and if applicable, details of automation tools used in the process. |  |
| Data collection process | 9 | Specify the methods used to collect data from reports, including how many reviewers collected data from each report, whether they worked independently, any processes for obtaining or confirming data from study investigators, and if applicable, details of automation tools used in the process. |  |
| Data items | 10a | List and define all outcomes for which data were sought. Specify whether all results that were compatible with each outcome domain in each study were sought (e.g. for all measures, time points, analyses), and if not, the methods used to decide which results to collect. |  |
|  | 10b | List and define all other variables for which data were sought (e.g. participant and intervention characteristics, funding sources). Describe any assumptions made about any missing or unclear information. |  |
| Study risk of bias assessment | 11 | Specify the methods used to assess risk of bias in the included studies, including details of the tool(s) used, how many reviewers assessed each study and whether they worked independently, and if applicable, details of automation tools used in the process. |  |
| Effect measures | 12 | Specify for each outcome the effect measure(s) (e.g. risk ratio, mean difference) used in the synthesis or presentation of results. |  |
| Synthesis methods | 13a | Describe the processes used to decide which studies were eligible for each synthesis (e.g. tabulating the study intervention characteristics and comparing against the planned groups for each synthesis (item #5)). |  |
|  | 13b | Describe any methods required to prepare the data for presentation or synthesis, such as handling of missing summary statistics, or data conversions. |  |
|  | 13c | Describe any methods used to tabulate or visually display results of individual studies and syntheses. |  |
|  | 13d | Describe any methods used to synthesize results and provide a rationale for the choice(s). If meta-analysis was performed, describe the model(s), method(s) to identify the presence and extent of statistical heterogeneity, and software package(s) used. |  |
|  | 13e | Describe any methods used to explore possible causes of heterogeneity among study results (e.g. subgroup analysis, meta-regression). |  |
|  | 13f | Describe any sensitivity analyses conducted to assess robustness of the synthesized results. |  |
| Reporting bias assessment | 14 | Describe any methods used to assess risk of bias due to missing results in a synthesis (arising from reporting biases). |  |
| Certainty assessment | 15 | Describe any methods used to assess certainty (or confidence) in the body of evidence for an outcome. |  |
| **RESULTS** | | |  |
| Study selection | 16a | Describe the results of the search and selection process, from the number of records identified in the search to the number of studies included in the review, ideally using a flow diagram. |  |
|  | 16b | Cite studies that might appear to meet the inclusion criteria, but which were excluded, and explain why they were excluded. |  |
| Study characteristics | 17 | Cite each included study and present its characteristics. |  |
| Risk of bias in studies | 18 | Present assessments of risk of bias for each included study. |  |
| Results of individual studies | 19 | For all outcomes, present, for each study: (a) summary statistics for each group (where appropriate) and (b) an effect estimate and its precision (e.g. confidence/credible interval), ideally using structured tables or plots. |  |
| Results of syntheses | 20a | For each synthesis, briefly summarise the characteristics and risk of bias among contributing studies. |  |
|  | 20b | Present results of all statistical syntheses conducted. If meta-analysis was done, present for each the summary estimate and its precision (e.g. confidence/credible interval) and measures of statistical heterogeneity. If comparing groups, describe the direction of the effect. |  |
|  | 20c | Present results of all investigations of possible causes of heterogeneity among study results. |  |
|  | 20d | Present results of all sensitivity analyses conducted to assess the robustness of the synthesized results. |  |
| Reporting biases | 21 | Present assessments of risk of bias due to missing results (arising from reporting biases) for each synthesis assessed. |  |
| Certainty of evidence | 22 | Present assessments of certainty (or confidence) in the body of evidence for each outcome assessed. |  |
| **DISCUSSION** | | |  |
| Discussion | 23a | Provide a general interpretation of the results in the context of other evidence. |  |
|  | 23b | Discuss any limitations of the evidence included in the review. |  |
|  | 23c | Discuss any limitations of the review processes used. |  |
|  | 23d | Discuss implications of the results for practice, policy, and future research. |  |
| **OTHER INFORMATION** | | |  |
| Registration and protocol | 24a | Provide registration information for the review, including register name and registration number, or state that the review was not registered. |  |
|  | 24b | Indicate where the review protocol can be accessed, or state that a protocol was not prepared. |  |
|  | 24c | Describe and explain any amendments to information provided at registration or in the protocol. |  |
| Support | 25 | Describe sources of financial or non-financial support for the review, and the role of the funders or sponsors in the review. |  |
| Competing interests | 26 | Declare any competing interests of review authors. |  |
| Availability of data, code and other materials | 27 | Report which of the following are publicly available and where they can be found: template data collection forms; data extracted from included studies; data used for all analyses; analytic code; any other materials used in the review. |  |

*From:*  Page MJ, McKenzie JE, Bossuyt PM, Boutron I, Hoffmann TC, Mulrow CD, et al. The PRISMA 2020 statement: an updated guideline for reporting systematic reviews. BMJ 2021;372:n71. doi: 10.1136/bmj.n71. This work is licensed under CC BY 4.0. To view a copy of this license, visit <https://creativecommons.org/licenses/by/4.0/>

| **PRISMA 2020 checklist item** | **Location where the item is reported** |
| --- | --- |
| **1. Title - Identify the report as a systematic review.** | **Title page:** “Vitamin D Status and Supplementation in Patients Undergoing Spine Surgery: A Systematic Review and Meta-analysis.” |
| **2. Abstract - Report the review according to the PRISMA 2020 for Abstracts checklist.** | **Abstract:** Background, Objective, Methods, Results, Conclusion, and Keywords. |
| **3. Rationale - Describe the rationale in the context of existing knowledge.** | **Introduction:** Burden of osteoporosis, biological role of vitamin D, relevance to spinal fusion, uncertainty and heterogeneity in the existing evidence. |
| **4. Objectives - Provide an explicit statement of the review objective or question.** | **End of Introduction :** “Accordingly, this systematic review and meta-analysis aimed to evaluate…” Primary and secondary outcomes are also identified. |
| **5. Eligibility criteria - Specify inclusion and exclusion criteria and how studies were grouped for synthesis.** | **Methods- Eligibility criteria :** Population, intervention or exposure, comparator, outcomes, study design, and exclusions. **Methods—Outcome definitions and analytical hierarchy, pp. 7–8:** direct supplementation analyses versus exploratory baseline-status or co-intervention analyses. |
| **6. Information sources - Specify all sources searched and the date of the last search.** | **Methods - Search strategy and information sources :** PubMed/MEDLINE, Scopus, Web of Science, Embase, and the Cochrane Library, searched from inception through **15 January 2026**. |
| **7. Search strategy - Present the full search strategies, filters, and limits.** | **Methods - Search strategy and information sources, pp. 4–5:** summary of search concepts, terms, language policy, and database adaptation. **Supplementary Material 1:** complete database-specific search strategies. |
| **8. Selection process - Describe how eligibility decisions were made, reviewer numbers, independence, and automation tools.** | **Methods-Study selection :** EndNote and Rayyan use, duplicate removal, four reviewers for title/abstract screening, two reviewers for full-text assessment, consensus resolution, and documentation of exclusion reasons. No automated eligibility-decision tool was used. |
| **9. Data collection process - Describe how data were collected, reviewer numbers, independence, investigator contact, and automation.** | **Methods. - Data extraction :** standardized form; two reviewers independently extracted data; disagreements were resolved through discussion and consensus. **Not applicable for investigator contact:** no process for obtaining additional data from study investigators was reported because the review used available aggregate published data. |
| **10a. Data items - Outcomes - List and define all outcomes and explain selection among measures or time points.** | **Methods-Outcomes; Data extraction ; Outcome definitions and analytical hierarchy :** postoperative VAS pain, ODI disability, fusion rate, time to fusion, pseudarthrosis, BMD, bone quality, implant complications, fractures, revision, quality of life, balance, inflammatory and muscle outcomes, and adverse events. . |
| **10b. Data items-Other variables - List other variables sought and assumptions concerning missing or unclear information.** | **Methods-Data extraction :** study, participant, surgical, medication, vitamin D, bone-health, treatment, and follow-up variables. **Not applicable for formal missing-data assumptions:** no imputation or assumptions for unavailable study-level information were reported; only available reported data were extracted. |
| **11. Study risk-of-bias assessment - Describe tools, reviewers, independence, and automation.** | **Methods-Risk-of-bias assessment :** design-specific JBI tools; two independent reviewers; item-level judgments; low, moderate, or high overall classifications; consensus resolution. No automated risk-of-bias tool was used. |
| **12. Effect measures - Specify the effect measure used for each outcome.** | **Methods-Outcome definitions and analytical hierarchy ; Data synthesis and quantitative analysis : mean** differences with 95% CIs for VAS and ODI; risk ratios with 95% CIs for fusion; lower MDs favor vitamin D-related groups, while RRs above 1 favor vitamin D-related treatment. |
| **13a. Synthesis eligibility - Explain how studies were selected for each synthesis.** | **Methods-Outcome definitions and analytical hierarchy :** direct supplementation studies were prioritized; baseline-status and major co-intervention comparisons were treated separately or as exploratory. **Methods-Data synthesis :** handling of relevant groups in multi-arm studies. |
| **13b. Data preparation - Describe handling of missing statistics, conversions, or other data preparation.** | **Methods-Outcome definitions and analytical hierarchy :** extraction of means, SDs, sample sizes, fusion events, and denominators at selected time points. **Not applicable for additional conversion or imputation:** no transformation of scales, imputation of missing SDs, or conversion of summary statistics was reported. |
| **13c. Tabulation and visual display - Describe methods used to tabulate or display results.** | Results are displayed in **Tables 1–3** , and **Figures 1–4** . Additional forest plots, subgroup plots, sensitivity analyses, influence diagnostics, and funnel plots are in **Supplementary Material 5**. Figure and table legends are present. |
| **13d. Statistical synthesis - Describe synthesis methods, model, heterogeneity methods, software, and rationale.** | **Methods-Data synthesis and quantitative analysis :** R version 4.5.1, random-effects models, restricted maximum likelihood, MDs, RRs, 95% CIs, Cochran’s Q, I², τ and τ², prediction intervals, and rationale based on anticipated clinical and methodological heterogeneity. |
| **13e. Exploration of heterogeneity - Describe subgroup analysis, meta-regression, or similar methods.** | **Methods-Subgroup, sensitivity, and influence analyses :** analyses according to direct supplementation versus clinically distinct bone-status/co-intervention comparisons, influence diagnostics, and study omission. **Not applicable for meta-regression:** too few studies were available for meaningful meta-regression. |
| **13f. Sensitivity analyses - Describe analyses used to assess robustness.** | **Methods-Subgroup, sensitivity, and influence analyses :** leave-one-out analyses, exclusion of Shi et al. from the intervention-focused pain analysis, exclusion of Haddadi et al. in an influence sensitivity analysis, supplementation-only fusion analysis, and alternative six-month Xu et al. fusion data. |
| **14. Reporting-bias assessment - Describe methods used to assess bias due to missing results.** | **Methods-Small-study effects :** funnel plots for VAS, ODI, overall fusion, and supplementation-only fusion; exploratory trim-and-fill analysis for VAS; explicit caution because fewer than 10 studies contributed to each outcome. |
| **15. Certainty assessment - Describe methods used to assess certainty of evidence.** | **Not applicable:** no formal GRADE or other certainty-of-evidence assessment was undertaken. |
| **16a. Study selection results - Report the search and selection process, preferably with a flow diagram.** | **Results-Study selection :** 3,793 records identified, 1,484 duplicates removed, 2,309 screened, 154 full texts assessed, 141 excluded, 13 studies included, and seven included in at least one synthesis. **Figure 1 :** PRISMA flow diagram. |
| **16b. Excluded studies - Cite apparently eligible excluded studies and explain exclusions.** | **Results-Study selection :** grouped exclusion reasons and counts. **Supplementary Material 2:** decisions and reasons for all 154 full-text reports, including the 141 excluded reports. Individual excluded studies are not cited separately in the main manuscript because the complete exclusion record is supplied in Supplementary Material 2. |
| **17. Study characteristics - Cite each included study and present its characteristics.** | **Results-Characteristics of the included studies ; Table 1 ; Table 2 .** All 13 included studies are cited as references **[21–33]**. |
| **18. Risk of bias in studies - Present the risk-of-bias assessment for each included study.** | **Results-Risk-of-bias assessment ; Table 3 :** study-level JBI summary. **Supplementary Material 4:** complete item-level assessments. |
| **19. Results of individual studies - Present study-level summary statistics and effect estimates.** | **Results-Narrative clinical outcomes ; Table 2 ; outcome-specific sections ; Figures 2–4 .** Study-specific estimates and CIs are shown for pooled outcomes; non-pooled outcomes are described narratively. |
| **20a. Characteristics and risk of bias among studies contributing to each synthesis.** | **Results-Characteristics of included studies ; Tables 1-2 ; outcome-specific synthesis sections ; Risk-of-bias assessment and Table 3 .** Characteristics and risk of bias are presented separately rather than repeated within every synthesis paragraph. |
| **20b. Results of statistical syntheses - Report pooled estimates, precision, heterogeneity, and direction.** | **Abstract ; Results-postoperative pain ; disability ; fusion rate ; Figures 2–4; Supplementary Material 5.** MDs or RRs, 95% CIs, p values, I², τ or τ², and prediction intervals where estimable are reported. |
| **20c. Results of investigations of heterogeneity.** | **Results-Pain, disability, and fusion analyses :** intervention-focused versus exploratory analyses, subgroup comparison for fusion, influence diagnostics, and changes in heterogeneity following study omission. **Supplementary Material 5.** |
| **20d. Results of sensitivity analyses.** | **Results - postoperative pain ; disability ; fusion rate :** omission of influential studies, leave-one-out results, supplementation-only analyses, and alternative follow-up-time analysis. **Supplementary Material 5.** |
| **21. Reporting biases - Present assessments of risk of bias due to missing results.** | **Results-Small-study effects ; Supplementary Material 5:** funnel plots and trim-and-fill findings. The manuscript appropriately states that no firm conclusion could be drawn because each analysis included fewer than 10 studies. |
| **22. Certainty of evidence - Present certainty assessments for each outcome.** | **Not applicable:** the review did not conduct GRADE or another formal certainty-of-evidence assessment. |
| **23a. Discussion-Interpretation - Interpret results in the context of other evidence.** | **Discussion :** Principal findings; Postoperative pain and disability; Fusion and bone-healing outcomes; Bone quality, implant stability, and fracture outcomes; Biological interpretation; Comparison with previous evidence. |
| **23b. Discussion-Limitations of included evidence.** | **Discussion-Limitations :** few studies, small samples, heterogeneity, observational designs, confounding, risk of bias, inconsistent fusion definitions, co-interventions, and imprecision. |
| **23c. Discussion-Limitations of review processes.** | **Discussion-Limitations :** limited studies per synthesis, inability to assess publication bias reliably, dependence on individual studies, heterogeneous outcome definitions and time points, and limitations of aggregate study-level data. |
| **23d. Discussion-Implications for practice, policy, and future research.** | **Discussion-Clinical implications ; Future directions ; Conclusion .** No specific policy recommendation is made because the evidence is insufficient. |
| **24a. Registration - Provide the register and registration number.** | **Methods-Protocol and reporting ; Statements and Declarations-Registration and reporting :** PROSPERO registration **CRD420261321165**. |
| **24b. Protocol access - State where the protocol can be accessed or that none was prepared.** | **Methods - Protocol and reporting ; Statements and Declarations-Registration and reporting .** The registered record is accessible through PROSPERO using **CRD420261321165**. |
| **24c. Protocol amendments - Describe and explain amendments to the protocol or registration.** | **Not applicable:** no amendments to the protocol or registered methods are described. To make this fully explicit, the manuscript may state: “No amendments were made to the registered protocol.” |
| **25. Support - Describe financial or non-financial support and the funder’s role.** | **Statements and Declarations-Funding :** no funds, grants, or other financial support were received. **Funder role not applicable because there was no funder.** |
| **26. Competing interests - Declare competing interests.** | **Statements and Declarations-Competing interests :** no relevant financial or non-financial interests were disclosed. |
| **27. Availability of data, code, and materials - State what is available and where.** | **Statements and Declarations - Data availability ; Supplementary material legends :** search strategies, full-text decisions, extraction dataset, item-level JBI assessments, and supplementary quantitative analyses are available in Supplementary Materials 1–6. |
