## Supplementary material for "Vitamin D Status and Supplementation in Patients Undergoing Spine Surgery: A Systematic Review and Meta-analysis": Tables

### Table 1. Characteristics of the 13 included studies.

| **Study** | **Country** | **Design** | **Ethics approval** |
| --- | --- | --- | --- |
| Haddadi et al. 2022 [21] | Iran | Double-blind randomized placebo-controlled trial | Yes |
| Hu et al. 2022 [22] | Taiwan | Randomized double-blind active-control trial | Yes |
| Ko et al. 2020 [23] | South Korea | Retrospective comparative cohort | Yes |
| Krasowska et al. 2019 [24] | Poland | Double-blind randomized controlled trial | Yes |
| Nekhlopochyn et al. 2025 [25] | Ukraine | Retrospective single-center cohort | Yes |
| Shafiee et al. 2021 [26] | Iran | Single-blind block-randomized clinical trial | Yes |
| Shi et al. 2024 [27] | China | Single-center retrospective cohort | Yes |
| Skrobot et al. 2019 [28] | Poland | Double-blind randomized controlled trial | Yes |
| Skrobot et al. 2020 [29] | Poland | Double-blind randomized controlled trial | Yes |
| Xu et al. 2014 [30] | China | Retrospective comparative cohort | Not reported |
| Dzik et al. 2019 [31] | Poland | Prospective comparative mechanistic analysis | Yes |
| Baumann et al. 2026 [32] | United States | Retrospective database cohort | Not reported |
| Li et al. 2025 [33] | Germany | Retrospective case-control study | Yes |

### Table 2. Vitamin D-related interventions or exposures, comparators, outcomes, and principal findings.

| **Study** | **Vitamin D-related intervention/exposure** | **Comparator** | **Baseline status** | **Outcomes** | **Principal findings** |
| --- | --- | --- | --- | --- | --- |
| Haddadi 2022 [21] | Oral vitamin D | Alendronate; control | All 25(OH)D >50 nmol/L | VAS, ODI, Modic change | No significant between-group differences; all groups improved from baseline. |
| Hu 2022 [22] | Vitamin D3 plus calcium | Calcium alone | Mixed sufficient/insufficient/deficient | Fusion, time to fusion, VAS, ODI | One-year fusion did not differ; vitamin D shortened time to fusion and improved selected 6-month VAS/ODI measures. |
| Ko 2020 [23] | Vitamin D supplementation | No supplementation | All deficient | ODI, RMDQ, SF-36 | ODI and SF-36 favored supplementation at 12 and 24 months; RMDQ did not differ. |
| Krasowska 2019 [24] | Preoperative vitamin D | Placebo | All insufficient | VAS, 25(OH)D, CRP, cytokines | 25(OH)D increased; pain did not clearly differ; selected inflammatory markers decreased after rehabilitation. |
| Nekhlopochyn 2025 [25] | Vitamin D3 plus K2 correction | No correction or no need for correction | Normal or deficient | 25(OH)D, CT bone quality, implant complications | Correction was associated with improved bone measures and fewer implant-related complications. |
| Shafiee 2021 [26] | Vitamin D | Alendronate; control | All 25(OH)D >10 ng/mL | Fusion, VAS | Fusion and between-group pain changes did not differ significantly. |
| Shi 2024 [27] | Calcium plus vitamin D, with/without denosumab | No treatment/normal bone status | Normal, osteopenic, or osteoporotic | Fusion, BMD, VAS, ODI, biomarkers | Outcomes differed by bone status and denosumab co-treatment; vitamin D’s independent effect could not be isolated. |
| Skrobot 2019 [28] | Vitamin D plus rehabilitation | Placebo plus rehabilitation | All sufficient | Limits of stability, postural stability, fall risk | Selected balance and stability measures improved more with vitamin D; fall risk did not clearly change. |
| Skrobot 2020 [29] | Vitamin D plus rehabilitation | Placebo plus rehabilitation | All sufficient | Balance, pressure distribution, fall risk | Several measures improved over time, with more pronounced selected improvements in the vitamin D group. |
| Xu 2014 [30] | 1,25-dihydroxyvitamin D3 | Control | Not reported | Radiographic fusion, ODI | Fusion and ODI favored treatment at 6 months and final follow-up. |
| Dzik 2019 [31] | Vitamin D supplementation/status groups | Sufficient and deficient placebo groups | Status defined by study | Muscle IGF-1, atrogin-1, mitochondrial markers | Vitamin D status and supplementation were associated with selected muscle metabolic and atrophy-signaling markers. |
| Baumann 2026 [32] | Vitamin D prescription | Other/no osteoporosis medication | Not reported | Postoperative sacral fracture | Vitamin D prescription was associated with lower sacral-fracture risk; causal inference is limited by the retrospective design. |
| Li 2025 [33] | Vitamin D3 dose categories | 0, <2,000, or ≥2,000 IU/day | Not reported | Pathological vertebral fragility, screw loosening, vertebral HU | Lower dose was associated with pathological fragility and lower HU; treatment was not randomized. |

### Table 3. Joanna Briggs Institute risk-of-bias summary.

| **Study** | **Checklist items** | **Yes** | **No** | **Unclear/partial** | **Overall risk** |
| --- | --- | --- | --- | --- | --- |
| Hu 2022 | 13 | 10 | 0 | 3 | Low |
| Krasowska 2019 | 13 | 9 | 0 | 4 | Moderate |
| Shafiee 2021 | 13 | 8 | 2 | 3 | Moderate |
| Haddadi 2022 | 13 | 10 | 0 | 3 | Moderate |
| Skrobot 2019 | 13 | 8 | 1 | 4 | High |
| Skrobot 2020 | 13 | 10 | 1 | 2 | Moderate |
| Dzik 2019 | 13 | 6 | 1 | 6 | High |
| Xu 2014 | 11 | 5 | 2 | 4 | Moderate |
| Nekhlopochyn 2025 | 11 | 9 | 1 | 1 | Low |
| Ko 2020 | 11 | 7 | 2 | 2 | Moderate |
| Shi 2024 | 11 | 6 | 3 | 2 | High |
| Baumann 2026 | 11 | 6 | 3 | 2 | Moderate |
| Li 2025 | 10 | 4 | 2 | 4 | Moderate |

*JBI, Joanna Briggs Institute. Item-level judgments are provided in Supplementary Material 4.*
